# Are effects of drinking in pregnancy modified by ADH1B and ALDH2 genotype? A longitudinal analysis in the Norwegian Mother, Father and Child Cohort Study (MoBa)

**DOI:** 10.64898/2026.10.20.26365709

**Authors:** Amanda M Hughes, Chloe Slaney, Kate Tilling, Paul Madley-Dowd, Alexandra Havdahl, George Davey Smith

## Abstract

**Background:** Recent evidence from a Japanese study suggests that adverse impacts of prenatal alcohol exposure (PAE) on child development are substantially modified by maternal genotype for rs671, a variant in the aldehyde dehydrogenase 2 gene (ALDH2) which impacts alcohol metabolism. This specific null variant is largely restricted to East Asian populations. Whether variants impacting alcohol metabolism in other ancestral groups could have similar effects is unclear.

**Methods:** Using data from the Norwegian Mother, Father, and Child Cohort Study (N=68,325 mother-child duos, data collected 2000-2011) we explored whether maternal drinking in early (1-15 weeks) or mid-pregnancy (13-30 weeks) was associated with children’s development. We considered the child’s language and motor skills at 18 and 36 months and the age when the child first walked unaided, as reported by mothers in questionnaires. Stratification by maternal and child genotypes linked to alcohol metabolism (rs1229984, rs886205 or rs440) examined effect modification.

**Results:** In minimally-adjusted models, children exposed to alcohol in early pregnancy (week 1-15) had slightly increased odds of delayed language skills at 18 months (OR=1.16 (95% CI: 1.01,1.33). This attenuated slightly with adjustment for confounders (OR=1.14 (95% CI: 0.99,1.31). Similar associations were seen for drinking in weeks 13-30, and there was little evidence of other harmful effects. In genetically-stratified models, there was little evidence that either the mother’s or child’s own genotype modified associations, but estimates for smaller genotype groups were imprecise.

**Conclusions:** Results suggest that in European ancestry populations, maternal or child rs1229984, rs886205 or rs440 genotype is unlikely to substantially modify consequences of prenatal alcohol exposure for early child development. Further work is required in European and other ancestry cohorts, including in countries where prevalence of drinking in pregnancy is higher.

## Background

In many European countries, public health advice is to abstain from alcohol completely while pregnant(1,2). This reflects conflicting evidence around risks of ‘light’ drinking, meaning there is no established ‘safe’ level of prenatal alcohol exposure (PAE)(3). Nevertheless, prenatal drinking in the WHO European Region is estimated at around 25%, the highest rate in the world(4).

Heavy drinking in pregnancy can cause developmental problems including delayed speech, language and motor development, cognitive impairment, difficulties with emotional and attention regulation even in children without facial or other physical features(5). In contrast, whether low to moderate PAE can impact children’s development is unresolved. Extensive research examining children’s cognitive, academic, socioemotional, and behavioural outcomes has reported detrimental, null, and even protective effects(6), with similarly conflicting results for birth outcomes(7,8). This may partly reflect inherent difficulties in studying PAE with survey data(6). First, alcohol consumption involves drinking frequency, typical and maximum units per occasion, and total units consumed. Surveys rarely capture all these dimensions, meaning studies have defined ‘low to moderate’ PAE inconsistently, and exposure windows vary greatly(6). Respondents often under-report consumption, which can bias estimates(9); this may be exacerbated by heightened stigma in pregnancy(10).

Second, associations may be confounded by risk factors for PAE which independently influence children’s development, including smoking(11), other substance use(12), poor mental health(12), and stressful life events, including domestic violence or abuse(13). PAE is also more common in unplanned pregnancies(14). Meanwhile, alcohol consumption shows very different social patterning to smoking. Women who drink during pregnancy, especially at ‘low’ or ‘moderate’ levels, typically have higher incomes and educational qualifications than abstainers(12,14–18). More generally, European health surveys report positive associations between alcohol consumption and higher socioeconomic status, especially for women(19,20). This complexity is under-recognised and poorly understood, perhaps because it challenges fundamental narratives about disadvantaged groups(21). A consequence is that PAE-exposed children often benefit from familial advantages which can bias associations with PAE(15,18).

Meanwhile, shared genetic factors could independently impact mothers’ alcohol consumption and children’s development(22). To circumvent confounding, researchers have used approaches which don’t rely on the assumption of no residual confounding of the exposure-outcome relationship, including instrumental variable analysis, sibling comparison, and negative control designs(23). Studies using instrumental variables for PAE suggests negative effects on children’s academic achievement(18); however, this design cannot describe nonlinear exposure effects, leaving the impact of ‘low to moderate’ PAE unclear. Sibling-comparison studies, which exploit differences in PAE between siblings to account for the shared family environment, suggest low-to-moderate PAE impairs children’s emotional and behavioural outcomes(22,24); negative control studies find less evidence of adverse effects(25,26).

Third, genetic factors linked to alcohol metabolism may influence risk of adverse effects following PAE. Alcohol metabolism has two principal stages: alcohol is oxidised to acetaldehyde by alcohol dehydrogenases (ADHs); resulting acetaldehyde is then oxidised to acetate by aldehyde dehydrogenase (ALDHs)(27). Genetic variants have been identified in European, East Asian and African ancestry populations which affect the efficiency of both stages. These variants increase the concentration of acetaldehyde in blood and organs after drinking, with unpleasant side effects (e.g., facial flushing, headache)(27). Consequently, they predict lower alcohol consumption and are protective against problematic drinking(28), and have therefore been used as instrumental variables(18,29).

A complexity is that these same variants may, whilst reducing alcohol consumption, also increase adverse effects when people do drink. Depending on genotype, different individuals are exposed to vastly different concentrations of acetaldehyde- a powerful carcinogen(30)- after consuming equivalent amounts of alcohol. A common East Asian variant (*ALDH2*2* / rs671 A allele) estimated to affect ∼540 million people(31) results in substantially reduced acetaldehyde clearance, and therefore higher blood acetaldehyde levels, after drinking (up to ∼6 times higher in heterozygotes, ∼20 times higher in homozygotes, compared to wildtype individuals(32)). Studies of oesophageal cancer show greater alcohol-associated risks for rs671 heterozygotes relative to wildtype individuals(33). Moreover, maternal rs671 could influence risk of developmental delay in children following PAE, by altering foetal exposure to acetaldehyde. A recent Japanese study (1727 mother-child duos) reported that children of rs671 heterozygote mothers who drank in pregnancy had increased risk of developmental delay across all five domains of the Ages and Stages Questionnaire (ASQ-3) at 36 months; for children of wildtype mothers, delay was only seen for communication(34). Follow-up work is needed, including in larger sample populations, and considering variants affecting acetaldehyde metabolism in other ancestral groups. Additionally, foetal genotype could modify consequences of PAE, by impacting foetal blood acetaldehyde; UK-based work suggests foetal variants can modify impact of PAE on IQ at age 8(35). We therefore use a large Norwegian pregnancy cohort to investigate associations between maternal alcohol consumption in pregnancy, maternal and foetal genotypes for alcohol and aldehyde metabolism, and children’s language and motor development in early childhood.

## Methods

### Participants

The Norwegian Mother, Father and Child Cohort Study (MoBa) is a population-based pregnancy cohort study conducted by the Norwegian Institute of Public Health(36). Participants were recruited from all over Norway from 1999-2008. The women consented to participation in 41% of the pregnancies. The cohort includes approximately 114,500 children, 95,200 mothers and 75,200 fathers. MoBa is regulated by the Norwegian Health Registry Act. The current study was approved by The Regional Committees for Medical and Health Research Ethics (2016/1702).

From all pregnancies in MoBa, we removed consent withdrawals, multiple births, and non-live births, leaving N=108,550 mother-child duos (Supplementary Figure 1). The last restriction could cause collider bias if PAE reduces probability of live births, but there was no strong evidence that in this cohort it did (Supplementary Methods). To explore associations of PAE and child development across all genotype groups, we restricted analysis to N=68,325 duos with complete data on variables likely to influence PAE (Supplementary Figure 2), before imputing missing values in PAE itself and outcomes. This follows recent evidence that multiple imputation with multiple incomplete variables may not be valid if incomplete variables are causes of missingness in other variables. In this case, restricting analysis to those with fully-observed early variables (subsample-MI) may be more likely to be valid, assuming the outcome is independent of selection into this subsample, conditional on analysis model variables(37). To explore associations stratified by maternal and child genotype, we further restricted to duos with genetic information(38) for mothers and children respectively (N=57,967 and N=49,776 duos) before imputing missing values in PAE and outcomes. Exclusion criteria and descriptive characteristics for each analysis is described in Supplementary Table 1-2, and specification of imputation models in Supplementary Table 3.

### Measures

#### PAE

In a questionnaire completed around 15 weeks’ gestation, MoBa mothers were asked about their alcohol consumption: ever (no/yes), in the three months before pregnancy, and during the pregnancy so far. For each period, mothers reported drinking frequency, units normally consumed, and frequency of ‘binge’ drinking (5+ units on one occasion). They also completed items from The Alcohol Use Disorders Identification Test (AUDIT)(39) including the units needed to feel an effect. Full details of all questions are available at https://www.fhi.no/en/ch/studies/moba/for-forskere-artikler/questionnaires-from-moba/. A later questionnaire, completed around 30 weeks’ gestation, asked about alcohol consumption in week 13-30 of the pregnancy. Frequency and heaviness of drinking in pregnancy were strongly left-skewed. We therefore derived binary measures (yes/no) of alcohol consumption during week 0-15 (based on the 15-week questionnaire), and week 13-30 (based on the 30-week questionnaire). Some responses were inconsistent (e.g., of 74,321 mothers who reported ‘never’ drinking at week 1-15, 2724 reported binge drinking in that period). We therefore coded participants to ‘yes’ if indicated by any response (Supplementary Methods). In additional analyses, we separated ‘light’ drinking (>0 but <5 units/occasion) from ‘binge’ drinking (5+ units on any occasion) in weeks 1-15 (this was not possible at weeks 13-30, given lower drinking prevalence). Item-level missingness meant PAE was missing for 6.4% and 9.8% respectively of respondents to the 15-week and 30-week questionnaires.

#### Child development

In questionnaires administered 18 and 36 months after the child’s birth, mothers completed questions relating to the child’s development, including adapted items from the Ages & Stages Questionnaire (ASQ). The original scale includes five developmental domains: communication, gross motor, fine motor, problem solving and social skills, with established cutoffs indicative of developmental delay(34). MoBa included a subset of ASQ items relating to communication and mixed fine and gross motor skills (Supplementary Methods). We follow previous work in this cohort(40,41), deriving summary scales for communication and mixed motor skills at 18 and 36 months. We apply standard ASQ cutoffs (2 standard deviations below the mean)(42) for binary measures of domain-specific delay. Additionally, we examined age (in months) when the child first walked unaided. This was reported at the 18 months and 36 months questionnaire; following previous work(43) we used the 18-month report where available, the 36-month report otherwise, and where both were available took the midpoint (Supplementary methods).

#### Genetic variants impacting alcohol and acetaldehyde metabolism

As possible genetic effect modifiers, we considered single nucleotide polymorphisms (SNPs) in key alcohol metabolism genes in European ancestral groups. For the alcohol to acetaldehyde stage, we used rs1229984, a non-synonymous SNP in the alcohol dehydrogenase 1B gene (*ADH1B*) which encodes the ADH1B enzyme. The minor allele leads to sharper increases in acetaldehyde concentration in blood and organs. Consequently, individuals with this variant drink less than wildtype individuals(18). Since only 0.01% of mothers had two copies of the acetaldehyde-increasing allele, rs1229984 genotype was coded as 0 vs. 1+ effect alleles (97.9% and 2.1% of mothers respectively). rs1229984 did not deviate from Hardy-Weinberg equilibrium (HWE p>0.05).

For the acetaldehyde to acetate stage, we considered SNPs in *ALDH2* or previously associated with ALDH2 expression in European ancestry populations, which were available in the MoBa data. We identified two regulatory variants, rs886205(44) and rs440(45). Other potentially relevant SNPs were either known to be in complete linkage disequilibrium with rs886205(46) or not available in MoBa(47,48). rs886205 and rs440 were coded as 0, 1, and 2 acetaldehyde-increasing alleles (rs886205: 66.6%, 29.9%, and 3.5% of mothers; rs440: 66.8%, 29.8%, 3.5% of mothers respectively). Unlike the East Asian variant rs671, effect of these SNPs on acetaldehyde clearance is modest(49). Therefore, to examine whether they could plausibly act as effect modifiers of PAE via efficiency of acetaldehyde clearance, we first examined their association with mother’s and father’s alcohol consumption before running stratified analyses. Neither SNP deviated from Hardy-Weinberg equilibrium (HWE p>0.05).

#### Covariates

Based on previous PAE literature, we identified potential confounders of associations between PAE and children’s development (Fig 1). From the Medical Birth Registry (MBRN), a national health registry containing information about all births in Norway, we took the child’s sex (male/female) and year of birth, the mother’s number of previous pregnancies, and the mother’s and father’s age. At the 15-week questionnaire, mothers reported whether the pregnancy was planned, their own income and the baby’s father’s annual income in broad categories, and their own and the baby’s father’s highest completed education. Where education reports were missing, we used administrative information on highest educational qualification in a later year (Supplementary methods). Smoking in pregnancy, categorized as no/yes, was derived from birth registry and questionnaire data (Supplementary Methods). Mother’s depressive/anxiety symptoms at pregnancy week 15 were derived from the 5-item Hopkins Checklist(50); whether the mother had experienced physical/sexual violence was coded as yes/no (Supplementary Methods). For ASQ outcomes, the child’s exact age (in months) when mothers completed the relevant questionnaire was included (Supplementary Methods). Because ancestry can confound genetic associations(51), models examining main effects of genotypes also included the first 20 principal components of ancestry (PCs) for the relevant individual, derived using the Psychgen pipeline(38,52).

**Figure 1:**
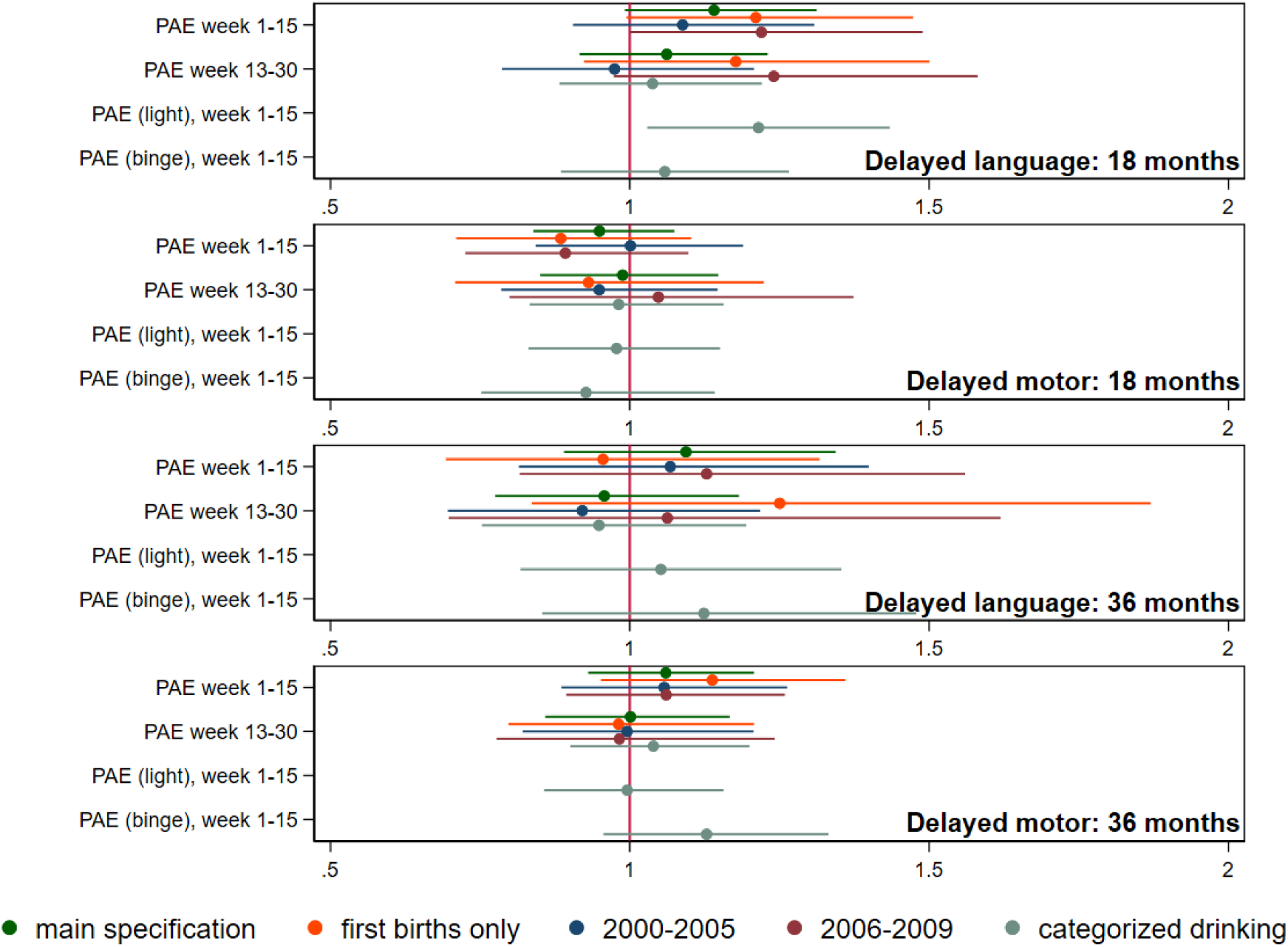
associations between prenatal alcohol exposure (PAE) and delayed language and motor skills at 18 and 36 months, main and alternative specifications (N=68,325) **Notes:** Associations expressed as odds ratios (ORs). Developmental delay is indicated by a score 2 S.D. below the mean on the relevant subscale of the modified Ages C Stages Questionnaire, based on information reported by mothers at 18 months and 36 months. ‘Light’ PAE defined as >0 but <5 units/occasion. ‘Binge’ PAE defined as 5+ units per occasion. All models adjust for: child’s sex, birth year, and exact at the 18-month or 36-month questionnaire, mother’s and father’s age, age squared, education and income, and the mother’s number of previous pregnancies, smoking and depressive/anxiety symptoms in pregnancy, experience of physical/sexual violence, and whether the pregnancy was planned.

### Statistical analysis

Main analyses explored associations of PAE with children’s development regardless of maternal or child genotype (N=68,325 duos, Analysis 1) while genotype-stratified analyses repeated these models within strata of maternal and child genotype (N=57,967 and N=49,776 duos respectively, Analysis 7-8). PAE at weeks 1-15 and 13-30 were included in models together; associations with PAE in each period are therefore mutually adjusted. To assess whether candidate SNPs were plausible effect modifiers of associations between PAE and children’s development, we examined associations of SNPs with mothers’ and fathers’ alcohol consumption before and during pregnancy (Analysis 2-3). This gave an indication of whether SNPs meaningfully impacted alcohol or acetaldehyde metabolism in this population, warranting genetically-stratified analyses. Only where there was evidence that SNPs influenced alcohol consumption did we proceed to analyses considering individual genotype groups (Analysis 4-9).

For each analysis, a separate imputation model was run (30 imputations), restricted to participants with complete information on factors influencing the analysis model’s exposure, and with genotype data for the mother, father and/or child as necessary, but otherwise maximising the available sample (Supplementary Tables 1-3). Each imputation model contained all model-specific exposures, outcomes and covariates and model-specific auxiliary variables (Supplementary Methods, Supplementary Table 1). All analysis models used robust standard errors.

#### Analysis 1: associations of PAE and children’s development across all genotype groups

Logistic regression was used to explore associations between PAE at weeks 1-15 and 13-30 and binary measures of communication and motor skills delay. Linear regression was used to explore associations between PAE and the age in months when the child first walked unaided. Associations with PAE at 1-15 and 13-30 weeks were mutually adjusted, for estimation of direct effects (i.e., effects of PAE at each timepoint, not via PAE at the other timepoint). All models adjusted for the child’s sex and year of birth, the mother’s and father’s age, age squared, educational qualifications and income, the mother’s number of previous pregnancies, smoking and depressive/anxiety symptoms in pregnancy, the mother’s experience of physical/sexual violence, whether the pregnancy was planned and, for ASQ outcomes, the child’s exact age in months when the outcome was reported (i.e., when the relevant questionnaire was completed).

We conducted sensitivity analyses. First, mothers may be less likely to drink during pregnancy following previous complicated births, potentially biasing associations with PAE among later-born children(25). We therefore followed previous work in restricting analyses to first births(8,25). Second, prevalence of drinking in pregnancy fell during the period of data collection, potentially changing the confounding structure and associations with children’s outcomes(8). We therefore stratified by year of child’s birth, splitting at the median (2000-2005 vs 2005-2009). Third, we examined associations with a categorized measure of PAE in weeks 0-15 (never/light/binge drinking). Separate imputation models were run for each sensitivity analysis, since these involved different exclusions or exposure specifications. We also reran main models using complete-case data.

#### Analysis 2-3: associations of genotypes with alcohol consumption in mothers and fathers

To examine whether rs1229984, rs886205 and rs440 were plausible genetic effect modifiers, we examined associations of mothers’ and fathers’ genotypes with their alcohol consumption before and during pregnancy (details: Supplementary Table 1). For mothers, we used linear regression to explore associations between the SNPs and pre-pregnancy drinking (the number of weekly drinking occasions, typical units/occasion, and frequency of binge drinking). We used logistic regression to explore associations with our main exposures: any drinking in week 1-15 and week 13-30 of pregnancy. For fathers, who completed different alcohol questions, we used linear regression to explore associations with weekly drinking occasions before and during the pregnancy. Analyses 2-3 were adjusted for the mother’s or father’s ancestry PCs only.

#### Analysis 4-c: associations of maternal, paternal, and child’s with children’s development

Where there was evidence that a SNP meaningfully impacted alcohol metabolism, we examined associations of that SNP in mothers and children with children’s development (details: Supplementary Table 1). As a negative control, we also examined associations with the SNP in fathers, since any association would reflect mechanisms independent of *in utero* exposure. Again, logistic and linear regression respectively were used for binary and continuous outcomes. Analyses 4-6 were adjusted for the relevant individual’s first 20 ancestry PCs, the child’s sex and year of birth and, for ASQ outcomes, the child’s age in months when the outcome was reported.

#### Analysis 7-8: associations of PAE with children’s development, stratified by maternal and child genotype

Where there was evidence that a SNP meaningfully impacted alcohol metabolism, we explored associations of PAE in weeks 1-15 and 13-30 with children’s development, stratified by maternal and child genotype (details: Supplementary Table 1). To allow associations to differ by genotype, separate imputation models were run for each genotype group.

#### Analysis S: associations of maternal and child genotype (mutually-adjusted) with children’s development, stratified by PAE

Correlation of genotypes within families means apparent effects of child genotype could be confounded by maternal genotype, and vice versa. We therefore also examined mutually-adjusted associations of maternal and child genotype with children’s development. Models were stratified by exposure, following stratified imputation (details: Supplementary Table 1).

## Results

### Descriptive characteristics

Over 99% of mothers reported having ever consumed alcohol; around 95% reported drinking in the 3 months before pregnancy. In contrast, only around 16% reported drinking in week 1-15 of pregnancy (Table 1), and 13% in weeks 13-30. Table 1 shows that, compared to mothers who did not drink in week 1-15, mothers who reported drinking were older and had markedly higher educational qualifications and higher income, consistent with previous studies (15–18). At the same time, mothers who drank in pregnancy were more often smokers, again consistent with previous evidence(16). Mothers for whom alcohol information was missing also differed.

**Table 1:**
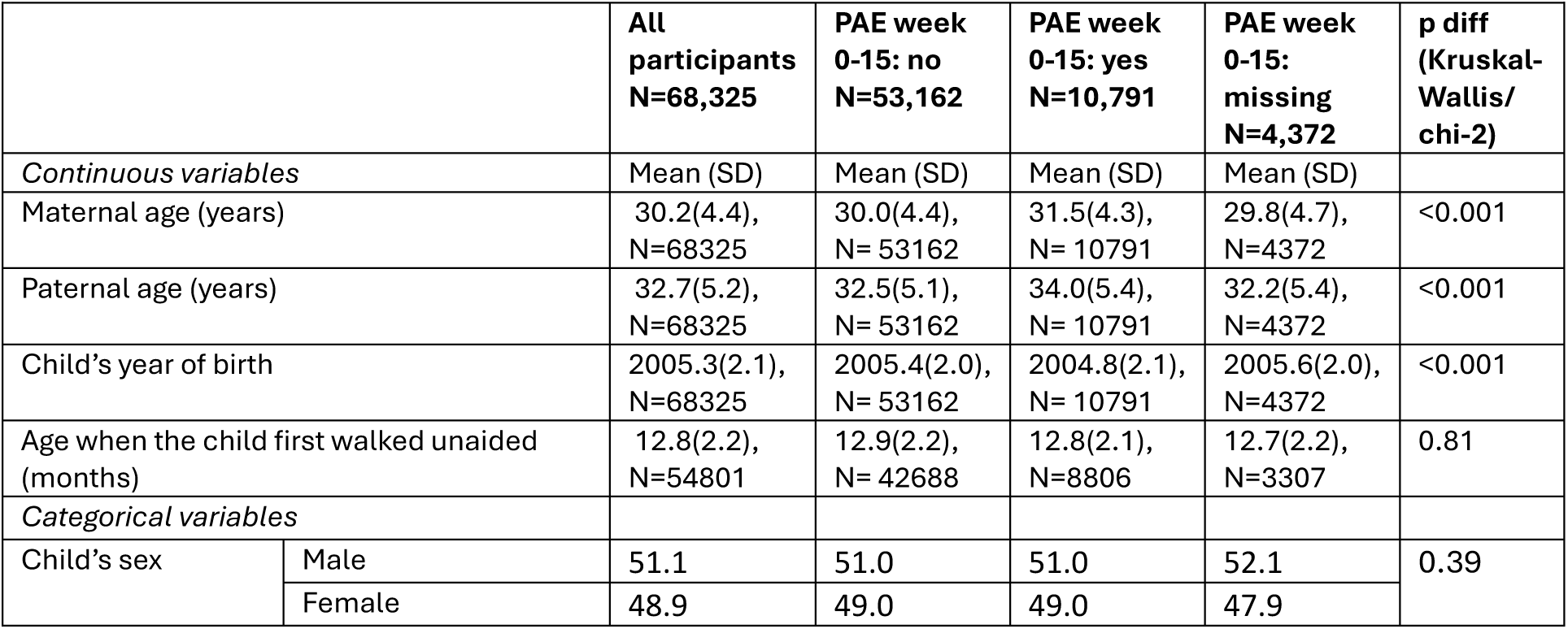
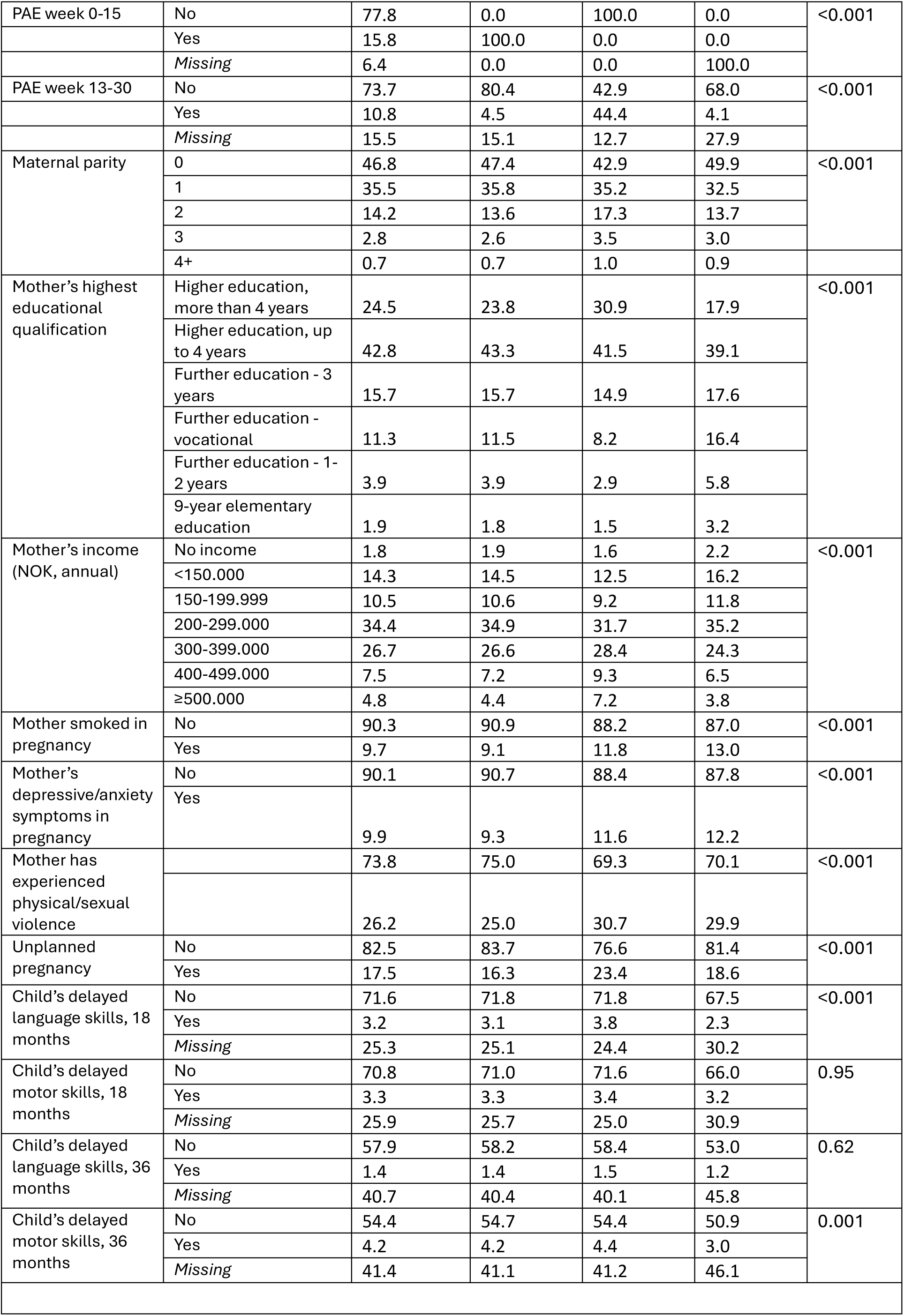
Descriptive characteristics of main analytical sample by PAE, pre-imputation.

Compared to mothers who did not drink, they were younger, with lower income, fewer qualifications, and fewer previous pregnancies. Supplementary Table 3 shows descriptive characteristics of the main analytical sample based on imputed data. In adjusted models, any drinking at week 1-15 and week 13-30 were positively associated with maternal education and income, but prevalence of ‘binge’ drinking at week 1-15 was similar across socioeconomic groups (Supplementary Table 5).

### Analysis 1: associations of PAE and child development across all genotypes

In models using imputed data and adjusted only for child’s sex, year of birth, and exact age at questionnaire completion, mothers who drank in week 1-15 of pregnancy were slightly more likely to report delayed language skills in their child at 18 months (OR=1.16 (95%CI: 1.01,1.33). This attenuated slightly with adjustment for further confounders (to OR=1.13 (95%CI: 0.99,1.31)); similar associations were seen for PAE at week 13-30 (Table 2). There was little evidence from any model that PAE in either period impacted language skills at 36 months or motor skills at either age (Table 2). In the least adjusted model, there was a small association between PAE at 13-30 weeks and age in months when the child began walking (mean difference:0.08 (95%CI:0.01,0.16), which attenuated with confounder adjustment (to 0.01, (95%CI: -0.07,0.08)).

**Table 2:** Associations of PAE and children’s development at different levels of adjustment.

| **Table 2: Associations of prenatal alcohol exposure (PAE) and all outcomes in the whole sample, different levels of adjustment** |  |  |  |  |
| --- | --- | --- | --- | --- |
|  | Week of alcohol exposure | Model 1* | Model 2: + mother's demographics and socioeconomic indicators† | Model 3: + all other covariates‡ |
|  |  | OR (95%CI) | OR (95%CI) | OR (95%CI) |
| Language delay at 18 months | 0-15 | 1.16(1.01,1.33) | 1.14(1.00,1.32) | 1.14(0.99,1.31) |
|  | 13-30 | 1.14(0.98,1.31) | 1.07(0.92,1.24) | 1.06(0.92,1.23) |
| Motor delay at 18 months | 0-15 | 0.97(0.86,1.10) | 0.95(0.84,1.07) | 0.95(0.84,1.07) |
|  | 13-30 | 1.07(0.92,1.24) | 0.99(0.85,1.15) | 0.99(0.85,1.15) |
| Language delay at 36 months | 0-15 | 1.09(0.89,1.33) | 1.12(0.91,1.37) | 1.09(0.89,1.34) |
|  | 13-30 | 0.95(0.77,1.17) | 0.97(0.79,1.20) | 0.96(0.78,1.18) |
| Motor delay at 36 months | 0-15 | 1.09(0.95,1.24) | 1.05(0.93,1.20) | 1.06(0.93,1.21) |
|  | 13-30 | 1.02(0.88,1.19) | 1.00(0.86,1.16) | 1.00(0.86,1.17) |
|  |  | Mean difference (95%CI) | Mean difference (95%CI) | Mean difference (95%CI) |
| Age at walking (months) | 0-15 | -0.02(-0.08,0.04) | -0.05(-0.11,0.01) | -0.04(-0.10,0.02) |
|  | 13-30 | 0.08(0.01,0.16) | 0.00(-0.07,0.08) | 0.01(-0.07,0.08) |
| *Adjusted for child's sex, year of birth, exact age in months at questionnaire completion. †Mother's and father's age, age squared, highest educational qualification and income, and the mother's number of previous pregnancies. ‡Mother's smoking, depressive/anxiety symptoms, experience of physical/sexual violence, whether the pregnancy was planned. |  |  |  |  |

Prevalence of PAE in weeks 1-15 and 13-30 reduced between 1999 and 2009 (Supplementary Table 5). Results were qualitatively similar when restricting to first-born children and in 1999-2005 vs 2005-2009 (Figures 1-2, Supplementary Table 6). Using a categorical exposure separating ‘light’ from ‘binge’ drinking in week 1-15, there was clearer evidence of associations between the child’s language skills at 18 months and ‘light’ drinking (OR=1.22, CI:1.03, 1.43) than ‘binge’ drinking (OR=1.06, CI:0.88,1.27) (Supplementary Table 6). Results based on complete-case data were qualitatively similar but estimates less precise (Supplementary Table 12).

**Figure 2:**
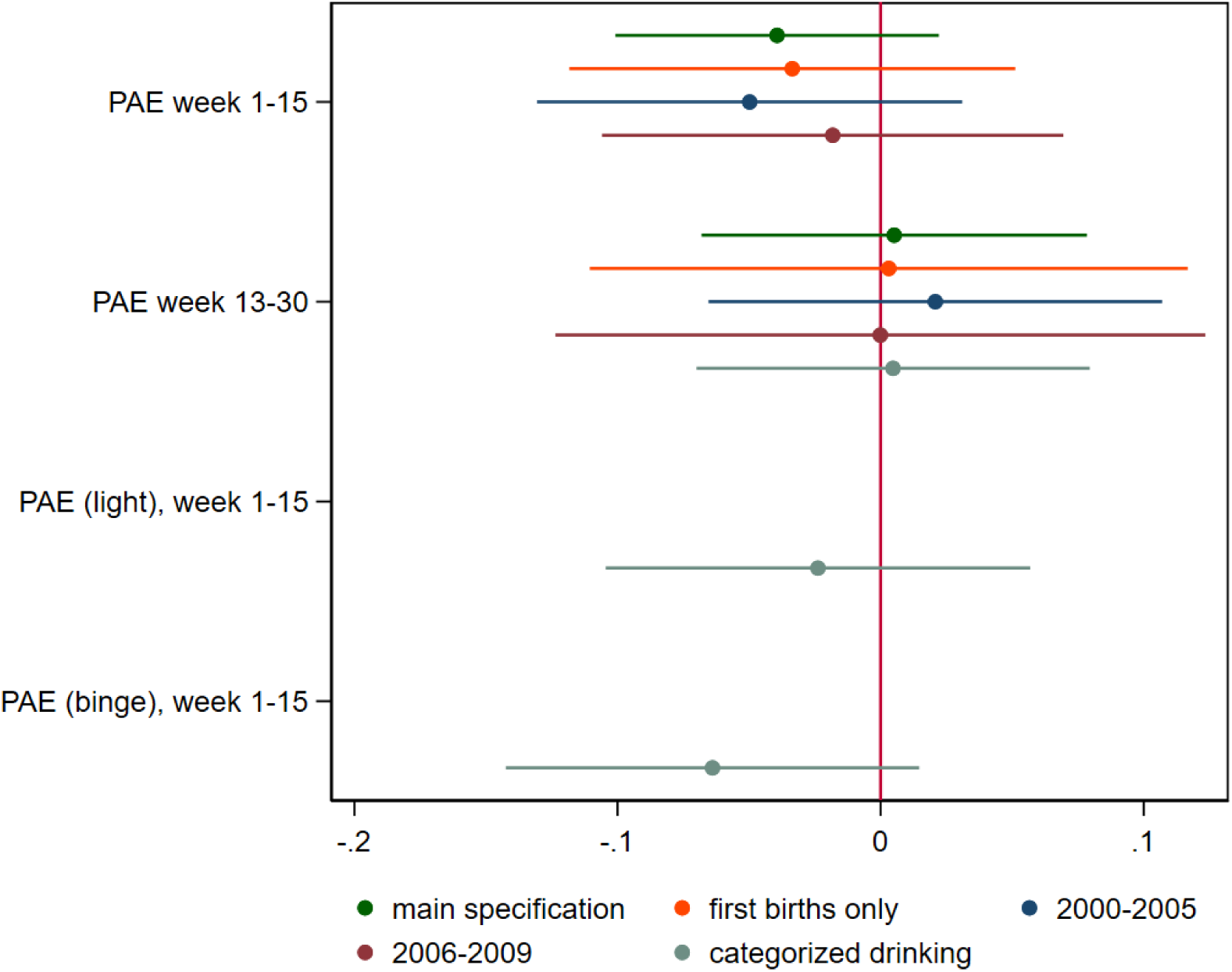
associations between prenatal alcohol exposure (PAE) and age when child first walked unaided, main and alternative specifications (N=68,325) **Notes:** Associations expressed as difference in months. ‘Light’ PAE defined as >0 but <5 units/occasion. ‘Binge’ PAE defined as 5+ units per occasion. All models adjust for: child’s sex and birth year, mother’s and father’s age, age squared, education and income, and the mother’s number of previous pregnancies, smoking and depressive/anxiety symptoms in pregnancy, experience of physical/sexual violence, and whether the pregnancy was planned.

### Analysis 2-3: association of SNPs with alcohol consumption

Adjusting for maternal ancestry PCs, maternal rs1229984 was consistently associated with drinking behaviour during and before pregnancy (Supplementary Table 7). Compared to mothers with 0 acetaldehyde-increasing allele, mothers with 1+ acetaldehyde-increasing alleles had fewer drinking occasions, typically drank fewer units per occasion, and had fewer ‘binge’ drinking occasions in the three months before pregnancy (all p<0.001). They were less likely to drink in week 1-15 (OR: 0.76, 95%CI:0.64,0.89) or week 13-30 of pregnancy (OR: 0.68, 95%CI:0.56,0.84). Using the categorical measure, point estimates suggested similar impact of rs1229984 on odds of ‘light’ and ‘binge’ drinking in weeks 1-15 (OR=0.69 (95%CI: 0.56, 0.86) and OR=0.87 (95%CI:0.67, 1.13)). Similarly, fathers with one or more acetaldehyde-increasing rs1229984 alleles drank less often before (-0.14 occasions/week, 95%CI: -0.20, -0.08) and during the pregnancy (by -0.11 occasions/week, 95%CI: -0.16, -0.05).

In contrast, there was little evidence that rs886205 or rs440 impacted drinking before or during pregnancy for mothers or fathers (Supplementary Table 7), suggesting these SNPs are unlikely to materially impact acetaldehyde levels after drinking in this population. We therefore dropped rs886205 and rs440 from analysis, focusing remaining analyses on rs1229984.

### Analysis 4-c: association of SNPs in mothers, fathers, and children with children’s development

In models adjusted for the relevant person’s ancestry PCs and the child’s sex, year of birth, and where relevant age at questionnaire completion, maternal and child rs1229984 genotype were both associated with slightly raised risk of delayed motor skills at 18 months (respectively, OR=1.31, 95%CI:1.00,1.72; OR=1.31, 95%CI:1.00,1.71). No associations were seen with other outcomes, or with paternal genotype (Supplementary Table 8).

### Analyses 7-8: association of PAE and children’s development, stratified by maternal and child genotype

In models stratified by maternal or child rs1229984 genotype estimates for the smaller genotype groups were imprecise, especially for ASQ outcomes, reflecting small case numbers (Supplementary Table 9). For all outcomes, there was little evidence of differential effects of PAE by maternal or child genotype, with substantially overlapping confidence intervals for all genotype groups (Figures 3-4, Supplementary Table 11).

**Figure 3:**
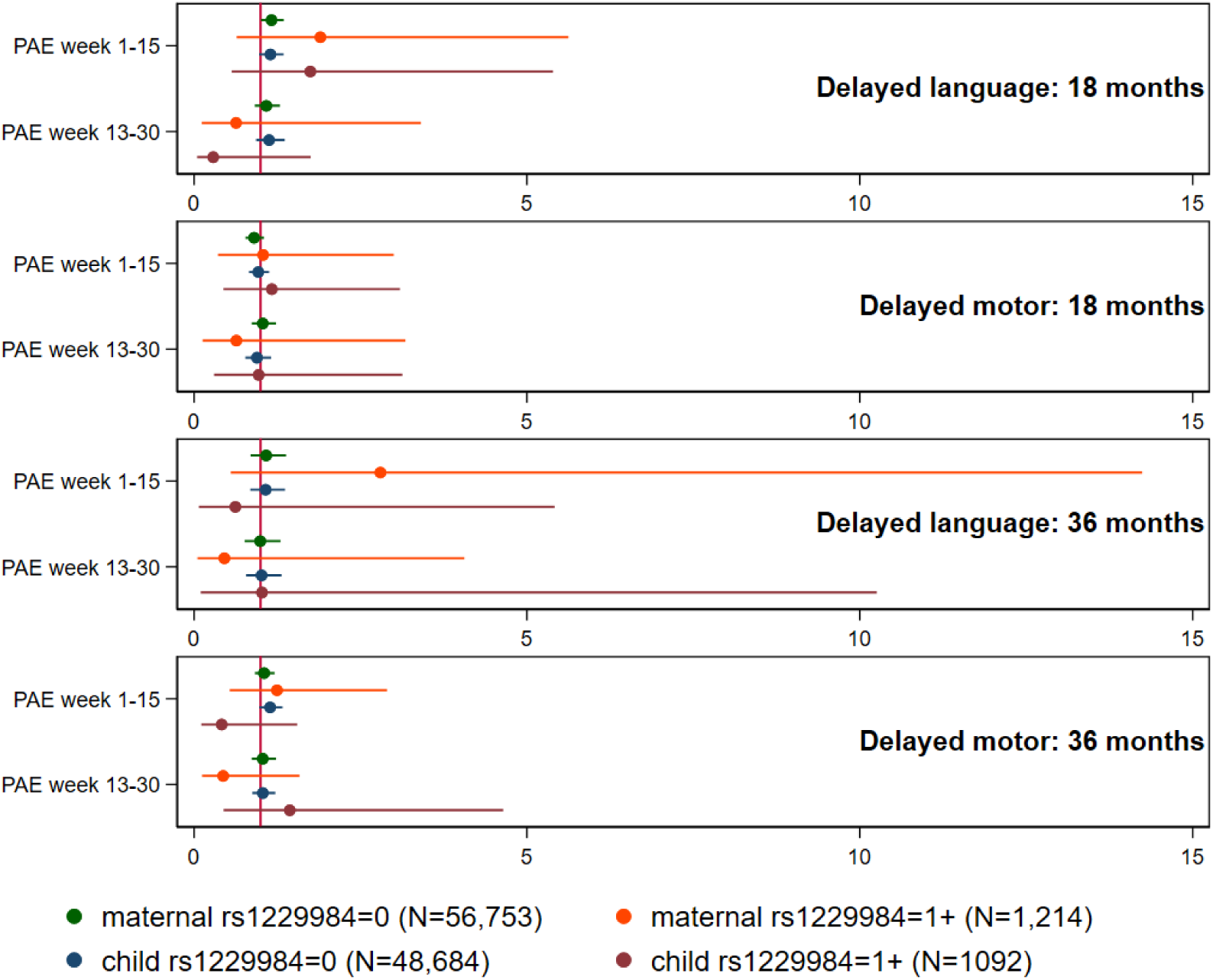
associations between prenatal alcohol exposure (PAE) and delayed language and motor skills at 18 and 36 months, stratified by maternal and child rs1229984 genotype. **Notes:** Associations expressed as odds ratios (ORs). Developmental delay is indicated by a score 2 S.D. below the mean on the relevant subscale of the modified Ages & Stages Questionnaire, based on information reported by mothers at 18 months and 36 months. All models adjust for: child’s sex, birth year, and exact at the 18-month or 36-month questionnaire, mother’s and father’s age, age squared, education and income, and the mother’s number of previous pregnancies, smoking and depressive/anxiety symptoms in pregnancy, experience of physical/sexual violence, and whether the pregnancy was planned.

**Figure 4:**
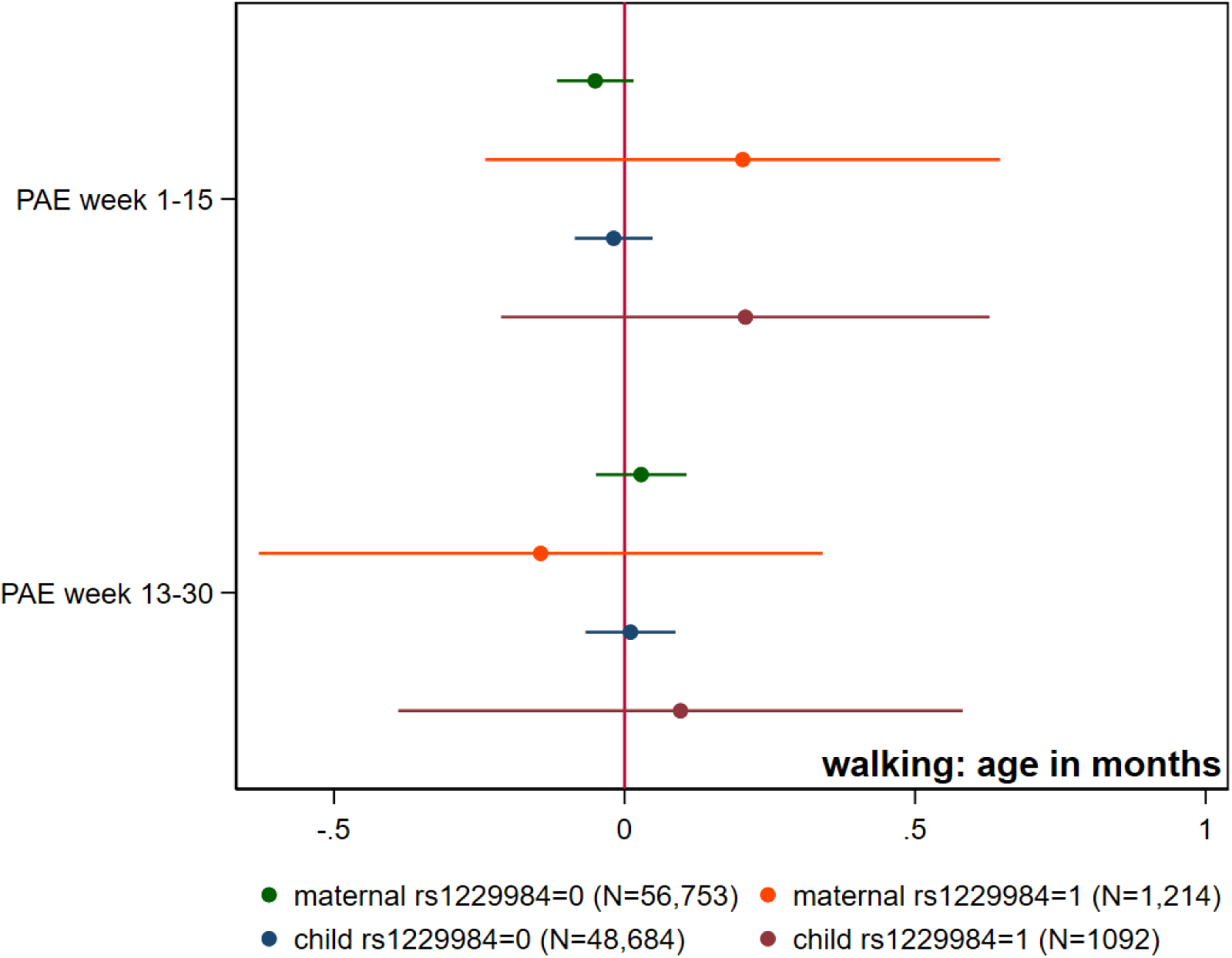
associations between prenatal alcohol exposure (PAE) and age when child first walked unaided, stratified by maternal and child rs1229984 genotype. **Notes:** Associations expressed as difference in months. All models adjust for: child’s sex and birth year, mother’s and father’s age, age squared, education and income, and the mother’s number of previous pregnancies, smoking and depressive/anxiety symptoms in pregnancy, experience of physical/sexual violence, and whether the pregnancy was planned.

### Analysis S: mutually-adjusted associations of maternal and child genotype with children’s development, stratified by PAE

Among children exposed to PAE, maternal rs1229984 genotype (1+ acetaldehyde-increasing alleles, vs. none) was associated with raised risk of delayed language skills at 36 months (OR=3.34, 95%CI:1.09,10.18), independently of the child’s genotype (Supplementary Table 10). No such associations were seen with other outcomes, or with the child’s own genotype. No associations were seen among unexposed children.

## Discussion

In a large Norwegian cohort, mothers who drank in early pregnancy (week 1-15) were slightly more likely than those who abstained to report delayed language skills in their child at 18 months. After confounder adjustment there was no strong evidence for an association, but small differences could not be ruled out. A similar pattern was seen for drinking in pregnancy week 13-30. No differences were seen for delayed language skills at 36 months, delayed motor skills at 18 or 36 months, or the age when the child first walked unaided. Since most mothers who drank in pregnancy did so only occasionally, these results are consistent with prior evidence on ‘light’ drinking, where studies often find limited evidence of impact. Analysis of ‘binge’ drinking in early pregnancy (5+ units on a single occasion) found little evidence for an impact, but numbers in this group were small, and in many cases may represent a single instance. For all outcomes, associations were broadly similar when restricted to first-born children, and for those born in 2000-2005 and in 2006-2009.

For all outcomes, there was little evidence that maternal genotype linked to alcohol metabolism modified associations. The *ADH1B* SNP rs1229984, which affects how quickly alcohol is metabolised to acetaldehyde, robustly predicted alcohol consumption in mothers and fathers before and during pregnancy, consistent with a substantial impact on blood acetaldehyde levels after drinking. Nevertheless, genetically-stratified models found little evidence that maternal or foetal rs1229984 moderates the impact of PAE on children’s early development. Similarly, direct associations of maternal and child rs1229984 with children’s outcomes showed no consistent pattern. This finding in a sample of 57967 mother-child duos is consistent with findings of an earlier study among 1727 mother-child pairs(34), suggesting limited prior evidence of moderation did not simply reflect lack of statistical power. Notably, we were also able to investigate moderation by foetal rs1229984 genotype. Our results contrast with those of an earlier UK-based study, where a polygenic index of four aldehyde dehydrogenase variants (including rs1229984) in children modified associations between PAE and children’s IQ at age 8(35). This may suggest subtle, longer-term impacts on development which may not be detectable with our binary measures at 18 and 36 months. Additionally, confidence intervals for smaller rs1229984 genotype groups in our study were wide, and smaller effect modifications cannot be ruled out. Lastly, if mothers with more acetaldehyde-increasing alleles who continue to drink during pregnancy typically drink less than those with fewer acetaldehyde-increasing alleles, this could balance greater impact at a given dosage, obscuring moderation. Our analysis suggested a similar effect of rs1229984 on odds of ‘light’ and ‘binge’ drinking (Supplementary Table 7), but more subtle differences in consumption would be difficult to detect.

Meanwhile, we did not investigate moderation by *ALDH2* variants because no candidate *ALDH2* SNP predicted alcohol consumption before or during pregnancy for mothers or fathers, indicating negligible effects on acetaldehyde clearance in this population. This contrasts with the *ALDH2* SNP common in East Asian populations (rs671), consistently shown to substantially impact drinking behaviour and implicated in a recent Japanese study as a modifier of the impact of PAE on multiple measures of children’s development(34). Together these findings suggest that, among European ancestry individuals, genetic variation in *ALDH2* may simply be less consequential, owing to smaller impacts of the relevant variants. Our candidate SNPs were identified from the previous literature, and additional variants linked to acetaldehyde clearance could have greater impacts on drinking behaviour and PAE. A genome-wide association study (GWAS) of blood acetaldehyde concentration following an alcohol challenge, or self-reported alcohol flushing, would allow combination of very small effects from many different SNPs. To our knowledge, no such GWAS exist.

### Strengths and limitations

A major strength of this analysis is its large sample size, with 68,325 mother-child duos for main analyses and up to 57,967 duos for genetically-stratified analyses. Drawing from a nationwide, multipurpose pregnancy cohort helped maximise representativeness. To minimise impact of missing data we applied multiple imputation, accounting for recent methodological work on minimising bias through careful sample definition(37). Alcohol consumption was reported anonymously via questionnaire, which reduces impact of social desirability bias relative to interviews(53), and prospectively. This is crucial, because bias can be exacerbated in retrospective reports made after children’s developmental problems emerge(6). Rich questionnaire data allowed separation of drinking in early and mid-pregnancy, and of ‘binge’ from lighter drinking. Sensitivity analyses, including year stratification and restriction to first-both children, strengthened robustness of conclusions. Notably, the large number of genotyped mothers and children enabled investigation of effect modification by maternal and children’s genotypes.

However, even in this large dataset, low prevalence of developmental delay meant estimates were imprecise, especially for the smaller genotype groups. Consequently, small main effects and interactions cannot be ruled out. A three-way interaction between PAE, maternal, and child’s own rs1228894 genotype is possible, but would require a substantially larger dataset to test. Analysis with genetic variants linked to acetaldehyde clearance was limited by a lack of *ALDH2* SNPs with detectable effects in this population; more work is needed to identify relevant variants in European ancestry populations, including GWAS. As with all self-reported alcohol data, there was missingness, inconsistencies, and possible underestimation. Given the socioeconomic patterning of PAE, residual confounding could have potentially masked harmful effects; however, adjustment for parental age, income and education made little difference, suggesting such impact is limited. Lastly, results from Norway-where alcohol consumption is lower than in most European countries, and sales strictly controlled and very heavily taxed(54)-may not generalize to other policy contexts. Further work in other European cohorts and beyond is now required.

## Conclusion

Results of genetic analyses in a large Norwegian birth cohort suggest that risks associated with drinking in pregnancy are unlikely to be substantially modified by maternal or foetal genetic factors linked to alcohol metabolism in European ancestry populations.

## Data availability

Data from the Norwegian Mother, Father and Child Cohort Study is managed by the Norwegian Institute of Public Health. Access requires approval from the Regional Committees for Medical and Health Research Ethics (REC), compliance with GDPR, and data owner approval.

Participant consent does not allow individual-level data storage in repositories or journals. Researchers seeking access for replication must apply via www.helsedata.no.

## Acknowledgements

We are grateful to all the participating families in Norway who take part in MoBa. For generating genomic data, we thank the Norwegian Institute of Public Health (NIPH), the HARVEST collaboration, the NORMENT Centre at the University of Oslo, the Center for Diabetes Research at the University of Bergen, deCODE Genetics, the Research Council of Norway, the South-Eastern and Western Norway Regional Health Authorities, the ERC AdG, Stiftelsen KG Jebsen, the Trond Mohn Foundation, and the Novo Nordisk Foundation.

## Supplementary information

### Supplementary Figures

**Supplementary Figure 1:**
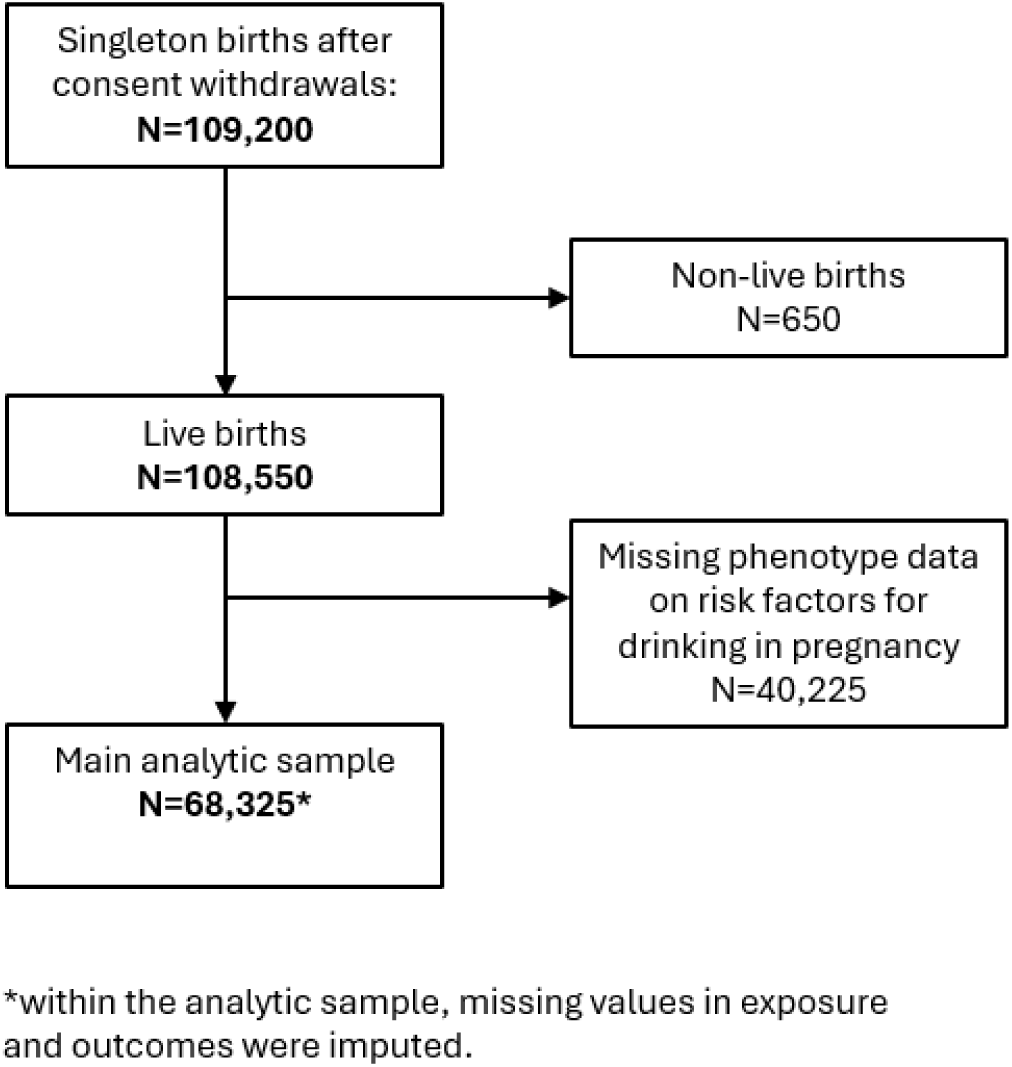
exclusions from the sample

**Supplementary Figure 2:**
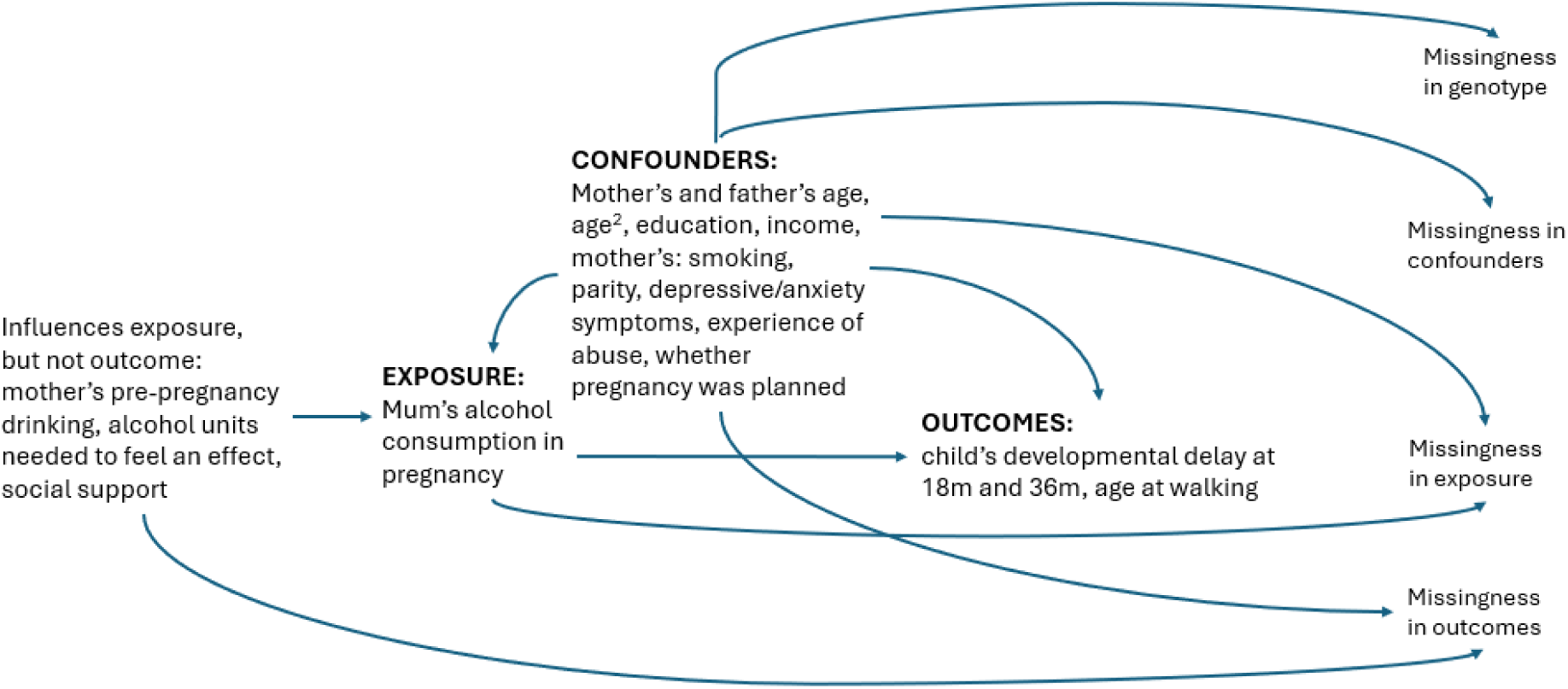
Directed Acyclic Graph of Key Variables

## Supplementary methods

### Sample exclusions

From all pregnancies in MoBa, we removed consent withdrawals, multiple births, and non-live births, before making further restrictions based on missing data (Supplementary Figure 1).

Restriction to live births could cause collider bias if PAE reduces probability of live births. Based on complete-case data, there was no strong evidence PAE impacted the chance of a live birth in this sample. In unadjusted models, ORs for PAE at weeks 1-15 and 13-30 respectively were: OR=1.04, 95%CI=0.83-1.30 (N=89,084) and OR=1.12, 95%CI=0.74-1.70 (N=80,330). In complete-case models adjusting for confounders (child’s sex and birth year, maternal and paternal age, income, education, maternal parity, smoking and depressive/anxiety symptoms in pregnancy, whether the pregnancy was planned), equivalent ORs for PAE at weeks 1-15 and 13-30 respectively were: OR=1.24, 95%CI=0.36-4.25 (N=6,532) and OR=1.38, 95%CI=0.15-12.48 (N=5,678).

### Coding of variables

#### Alcohol items at 15 weeks’ questionnaire

The main exposure was defined as 0 (no drinking), 1 (some drinking), and 2 (binge drinking), in the first 15 weeks. To derive this, we used self-reported information from the 15 weeks’ gestation questionnaire on several dimensions of drinking behaviour since becoming pregnant. We drew on: drinking frequency (never; less than once a month; 1-3 times a month; once a week; 2-3 times a week; 4-5 times a week; 6-7 times a week), units per occasion (less than 1; 1-2; 3-4; 5-6; 7-9; 10 or more) and frequency of drinking 5 units or more (never; less than once a month; 1-3 times a month; once a week; several times per week). All values were initially set to missing. First, we considered drinking frequency: mothers were assigned a 0 if they reported ‘never’ drinking since becoming pregnant, and a 1 if they reported any other frequency. Second, we considered units per occasion. Regardless of the reported frequency, mothers were assigned a 1 if they reported any units per occasion greater than the minimum (<1). Mothers still with a missing value were assigned a 0 if they reported the minimum (<1). Third, regardless of answers to previous questions, we coded the exposure to 2 (‘binge’) for mothers who reported having ever drank 5 units or more since becoming pregnant. Finally, we set remaining missing values to 0 for mothers who had answered ‘no’ to the question ‘have you ever consumed alcohol?’ Full details of all questions are available at https://www.fhi.no/en/ch/studies/moba/for-forskere-artikler/questionnaires-from-moba/.

#### Child developmental outcomes

Communication and motor skills at 18 and 36 months were based on mother-reported information from questionnaires. The original scale includes five developmental domains: communication, gross motor, fine motor, problem solving and social skills, with established cutoffs indicative of developmental delay(34). MoBa included a subset of ASQ items relating to communication and mixed fine and gross motor skills, listed below. For each item, mothers could respond: 1=*‘Yes, often’,* 2=‘*Sometimes’*, or *3=*‘*Not yet’*. For each set of items, mothers’ responses were summed and reverse-coded so that higher scores indicated lower skill. We applied standard ASQ cutoffs of 2 standard deviations below the mean(42) to derive binary measures of domain-specific delay.

#### Communication skills at 18 months

1. When you ask him/her, does your child go into another room to find a familiar toy or object? (You might ask, “Where is your ball?”, or say, “Bring me your coat” or “Go get your blanket”).

2. Does your child say eight or more words in addition to “mama” and “dada”?

3. Without showing him/her first, does your child point to the correct picture when you say, “Show me the kitty” or ask, “Where is the dog”?

#### Communication skills at 3c months

4. Without showing him/her first, does your child point to the correct picture when you say, “Where is the cat” or “Where is the dog”? Your child must only point at the correct picture

5. When you ask your child to point to his/her eyes, nose, hair, feet, ears, and so forth, does he/she correctly point to at least seven body parts? (The child can point to parts of himself/herself, you, or a doll.)

6. Does your child make sentences that are three or four words long?

7. Without giving him/her help by pointing or using gestures, ask your child to “Put the shoe on the table” and “Put the book under the chair”. Does your child carry out both of these directions correctly?

8. When looking at a picture book, does your child tell you what is happening or what action is taking place in the picture? (For example, “Barking”, “Running”, “Eating” and “Crying”?) You may ask, “What is the dog (or boy) doing?”

9. Can your child tell you at least two things about an object he/she is familiar with? If you say, for example, “Tell me about your ball”, will your child answer by saying something like “It is round, I can throw it, it is big”?

#### Motor skills at 18 months

1. Can your child walk well and seldom fall?
2. Does your child walk down stairs if you hold onto one of his/her hands?
3. Does your child throw a small ball or toy with a forward arm motion? (If he/she simply drops the ball, enter a cross under “Not yet”)
4. Does your child stack a small block or toy on top of another one? (For example, small boxes or toys about 3 cm in size)
5. Does your child turn the pages of a book by himself/herself? (He/she may turn more than one page at a time.)
6. Does your child move around by walking, rather than by crawling on his/her hands and knees?

#### Motor skills at 3c months

1. Without holding onto anything for support, does your child kick a ball by swinging his/her leg forward?
2. Can your child catch a large ball with both hands?
3. When drawing, does your child hold a pencil, crayon, or pen between his/her fingers and thumb like an adult does?
4. Can your child undo one or more buttons?

#### Age at walking

We also examined age (in months) at which the child first walked unaided. This was reported at both the 18 months and 36 months questionnaire. Following previous work in this cohort(35), we used the 18-month report where available, the 36-month report otherwise, and where both were available took the midpoint. Implausible values (i.e., children walking before 6 months(43), or later than the child’s age when the questionnaire was completed) were set to missing. For a very small number of cases (∼0.4% of the sample) the last available report was that the child was ‘not yet walking’; these were set to the following month.

#### Covariates

As part of the 15 weeks’ gestation questionnaire, mothers reported their own and the baby’s father’s annual income, choosing from: no income; <150,000 NOK; 150,000-199,999 NOK; 200,000-299,999 NOK; 300,000-399,999 NOK; 400,000-499,999 NOK; 500,000 NOK or above.

Mothers also reported their own and the baby’s father’s highest completed education, choosing from: 9-year secondary school; 1-2 year high school; vocational high school; 3-year high school general studies, junior college; regional technical college, 4-year university degree (bachelor’s degree, nurse, teacher, engineer); university, technical college, more than 4 years (master’s degree, medical doctor, PhD). For a small proportion of cases with missing questionnaire data on education (2% of mothers, 5% of fathers), we used administrative data on highest education completed in a later year. Mother’s smoking in early pregnancy (yes/no) was derived using information from the birth registry (83% of cases), substituting in a self-report from the 15 weeks’ gestation questionnaire where birth registry data was missing (17% of cases). For a further 0.02% of participants lacking both indicators, we used smoking in late pregnancy (from the birth registry file, or 30 weeks’ gestation questionnaire). Mother’s depressive/anxiety symptoms in pregnancy were derived from a 5-item version of the Hopkins Checklist, self-completed as part of the 15-week questionnaire. Due to strong skew, this was dichotomized using a cutoff of ≥1.80, previously found to have the best balance of sensitivity and specificity in a general population sample of Norwegian women(50). Mother’s history of physical/sexual violence was derived from questions completed in the 15-week questionnaire. Individual questions asked about physical and sexual violence i) during the pregnancy, ii) in the six months before, and iii) earlier (all yes/no). Due to small numbers, these were combined into a single binary measure. The child’s exact age in months when mothers completed the 18-month and 36-month questionnaires was available for almost all returned questionnaires (98.2%, 97.0% respectively). Where this was missing, we took the child’s age in months at the midpoint between the date the questionnaire was sent out by MoBa and the date it was returned. Where the questionnaire had not been returned, we assigned the median value, allowing us to include these as regular variables in imputation models.

#### Multiple imputation

Multiple imputation (m=30) by chained equations was performed using Stata’s mi impute (details in Supplementary Table 3). Binary variables were imputed with logistic regression, categorical variables with multinomial logistic regression, and continuous variables with predictive mean matching (knn=10). Imputation models contained all exposures, outcomes, and confounders used in analysis models. As auxiliary variables for alcohol exposures, we include additional factors which could influence the mother’s alcohol consumption in pregnancy, including: pre-pregnancy alcohol consumption, the mother’s pre-pregnancy body mass index, whether she exercised regularly, and social support. For analyses stratified by maternal or child genotype, imputation was restricted to those with complete data on the stratifying variable, and imputation models run separately within each stratum.

---

**Supplementary Table 1:** Sample definition for all analyses.

| **Supplementary Table 1: Sample definition for all analyses** |  |  |  |
| --- | --- | --- | --- |
| **Analysis** | **N** | **Restricted to those with complete:** | **Covariates in analysis model** |
| **Analysis 1:** Effect of PAE at week 1-15 and week 13-30 on offspring outcomes | 68325 | Confounders<br>Other influences on PAE* | Confounders, PAE at the other timepoint<sup>‡</sup> |
| **Analysis 2:** Association of ADH1B and ALDH2 with maternal alcohol intake before and during pregnancy. | 57967 | Maternal genotype<br>Confounders*<br>Other influences on PAE* | Maternal ancestry PCs |
| **Analysis 3:** Association of ADH1B and ALDH2 with paternal alcohol intake before and during pregnancy. | 48801 | Paternal genotype<br>Influences on father's drinking*<sup>†</sup> | Paternal ancestry PCs |
| **Analysis 4:** Association of maternal ADH1B with offspring outcomes | 70242 | Maternal genotype<br>Confounders<sup>‡</sup> | Maternal ancestry PCs |
| **Analysis 5:** Association of paternal ADH1B with offspring outcomes | 50176 | Paternal genotype<br>Confounders<sup>‡</sup> | Paternal ancestry PCs |
| **Analysis 6:** Association of offspring ADH1B with outcomes | 60215 | Offspring genotype<br>Confounders<sup>‡</sup> | Offspring ancestry PCs |
| **Analysis 7:** Effect of PAE on offspring outcomes, stratified by maternal ADH1B | 57967 | Confounders<br>Other influences on PAE*<br>Maternal genotype | Confounders, PAE at the other timepoint<sup>‡</sup> |
| **Analysis 8:** Effect of PAE on offspring outcomes, stratified by child ADH1B | 49776 | Confounders<br>Other influences on PAE*<br>Offspring genotype | Confounders, PAE at the other timepoint<sup>‡</sup> |
| **Analysis 9:** Joint (mutually-adjusted) analysis of maternal and offspring genotype with offspring outcomes, stratified by PAE. | 41758 (no PAE at week 1-15) + 7923 (PAE at week 1-15) | Confounders**<br>Maternal genotype<br>Offspring genotype<br>PAE (since stratifying on this) | The other person's genotype, maternal and offspring ancestry PCs |
| *Not included in analysis model, but used as auxiliary variables in imputation of alcohol consumption to reduce the impact of data MNAR. <sup>†</sup>Father's and mother's age, age<sup>2</sup>, educational qualifications and income, father's smoking, BMI, depressive symptoms. <sup>‡</sup>Not included in analysis model, but used as auxiliary variables in imputation of offspring outcomes to reduce the impact of data MNAR. <sup>§</sup>Meaning direct effects of PAE at each timepoint estimated. |  |  |  |

**Supplementary Table 2:** Descriptive characteristics of analytic sample for all analyses (pre-imputation)

|  |  |  |  |  |  |  |  |  |  |  |
| --- | --- | --- | --- | --- | --- | --- | --- | --- | --- | --- |
|  |  | Analysis 1<br>N=68,325 | Analysis 2<br>N=57967 | Analysis 3<br>N=48801 | Analysis 4<br>N=70242 | Analysis 5<br>N=50176 | Analysis 6<br>N=60215 | Analysis 7<br>N=57967 | Analysis 8<br>N=49776 | Analysis 9<br>N=49681 |
| *Continuous variables* |  | Mean (SD), N<br>non-missing | Mean (SD), N<br>non-missing | Mean (SD), N<br>non-missing | Mean (SD),<br>N non-missing | Mean (SD), N<br>non-missing | Mean (SD), N<br>non-missing | Mean (SD), N<br>non-missing | Mean (SD), N<br>non-missing |  |
| Maternal age (years) |  | 30.2(4.4),<br>N=68325 | 30.3(4.4),<br>N=57967 | 30.2(4.4),<br>N=48801 | 30.3(4.5),<br>N=70242 | 30.2(4.4),<br>N=50176 | 30.2(4.4),<br>N=60215 | 30.3(4.4),<br>N=57967 | 30.2(4.4),<br>N=49776 | 30.3(4.4),<br>N=49681 |
| Paternal age (years) |  | 32.7(5.2),<br>N=68325 | 32.7(5.2),<br>N=57967 | 32.6(5.1),<br>N=48801 | 32.7(5.3),<br>N=70242 | 32.6(5.1),<br>N=50176 | 32.7(5.2),<br>N=60215 | 32.7(5.2),<br>N=57967 | 32.7(5.1),<br>N=49776 | 32.7(5.2),<br>N=49681 |
| Child's year of birth |  | 2005.3(2.1),<br>N=68325 | 2005.3(2.1),<br>N=57967 | 2005.6(1.9),<br>N=48801 | 2005.2(2.1),<br>N=70242 | 2005.5(1.9),<br>N=50176 | 2005.2(2.1),<br>N=60215 | 2005.3(2.1),<br>N=57967 | 2005.3(2.1),<br>N=49776 | 2005.2(2.1),<br>N=49681 |
| Child's age at walking (months) |  | 12.8(2.2),<br>N=54801 | 12.8(2.2),<br>N=46980 | 12.9(2.3),<br>N=39280 | 12.8(2.2),<br>N=56259 | 12.9(2.3),<br>N=40247 | 12.8(2.1),<br>N=48863 | 12.8(2.2),<br>N=46980 | 12.8(2.1),<br>N=40840 | 12.8(2.2),<br>N=40528 |
| *Categorical variables* |  | % | % | % | % | % | % | % | % |  |
| Child's sex | Male | 51.1 | 51.1 | 51.1 | 51.0 | 51.1 | 51.1 | 51.1 | 51.2 | 51.0 |
|  | Female | 48.9 | 48.9 | 48.9 | 49.0 | 48.9 | 48.9 | 48.9 | 48.8 | 49.0 |
| Maternal parity | 0 | 46.8 | 46.5 | 48.1 | 44.8 | 47.5 | 44.8 | 46.5 | 46.4 | 44.6 |
|  | 1 | 35.5 | 35.8 | 35.0 | 36.2 | 35.5 | 36.1 | 35.8 | 35.8 | 36.4 |
|  | 2 | 14.2 | 14.2 | 13.6 | 15.0 | 13.6 | 15.1 | 14.2 | 14.4 | 15.1 |
|  | 3 | 2.8 | 2.8 | 2.6 | 3.1 | 2.7 | 3.1 | 2.8 | 2.7 | 3.1 |
|  | 4+ | 0.7 | 0.7 | 0.7 | 0.9 | 0.7 | 0.9 | 0.7 | 0.7 | 0.9 |
| Mother's highest<br>educational<br>qualification | Higher education,<br>more than 4 years | 24.5 | 24.4 | 24.5 | 23.3 | 24.6 | 23.2 | 24.4 | 24.2 | 23.5 |
|  | Higher education,<br>up to 4 years | 42.8 | 43.1 | 43.1 | 42.6 | 43.3 | 42.9 | 43.1 | 43.4 | 43.4 |
|  | Further education -<br>3 years | 15.7 | 15.6 | 15.0 | 15.8 | 15.0 | 15.8 | 15.6 | 15.6 | 15.6 |
|  | Further education -<br>vocational | 11.3 | 11.3 | 11.6 | 12.0 | 11.4 | 11.9 | 11.3 | 11.3 | 11.6 |
|  | Further education -<br>1-2 years | 3.9 | 3.8 | 3.9 | 4.3 | 3.8 | 4.2 | 3.8 | 3.7 | 4.0 |
|  | 9-year elementary<br>education | 1.9 | 1.8 | 1.9 | 2.0 | 1.9 | 2.0 | 1.8 | 1.7 | 1.9 |
| Mother's income<br>(NOK, annual) | No income | 1.8 | 1.7 | 1.9 | 1.9 | 1.8 | 1.8 | 1.7 | 1.7 | 1.8 |
|  | <150.000 | 14.3 | 14.0 | 14.0 | 14.9 | 13.8 | 14.9 | 14.0 | 14.0 | 14.7 |
|  | 150-199.999 | 10.5 | 10.5 | 10.1 | 11.0 | 10.1 | 11.1 | 10.5 | 10.6 | 11.0 |
|  | 200-299.000 | 34.4 | 34.7 | 34.2 | 34.8 | 34.4 | 35.3 | 34.7 | 35.2 | 35.3 |
|  | 300-399.000 | 26.7 | 26.7 | 27.6 | 25.7 | 27.5 | 25.6 | 26.7 | 26.6 | 25.8 |
|  | 400-499.000 | 7.5 | 7.5 | 7.5 | 7.0 | 7.6 | 7.0 | 7.5 | 7.4 | 7.1 |
|  | ≥500.000 | 4.8 | 4.8 | 4.8 | 4.5 | 4.8 | 4.3 | 4.8 | 4.6 | 4.4 |
| PAE week 1-15 | No | 77.8 | 77.9 | 78.9 | 77.7 | 78.8 | 77.9 | 77.9 | 78.1 | 84.1 |
|  | Yes | 15.8 | 15.9 | 14.0 | 14.9 | 14.4 | 14.8 | 15.9 | 15.6 | 15.9 |
|  | Missing | 6.4 | 6.3 | 7.1 | 7.4 | 6.9 | 7.4 | 6.3 | 6.3 |  |
| PAE week 13-30 | No | 73.7 | 74.5 | 75.4 | 73.5 | 74.9 | 74.8 | 74.5 | 75.8 | 75.5 |
|  | Yes | 10.8 | 10.9 | 9.4 | 10.4 | 9.8 | 10.4 | 10.9 | 10.9 | 10.8 |
|  | Missing | 15.5 | 14.5 | 15.3 | 16.1 | 15.3 | 14.7 | 14.5 | 13.3 | 13.7 |
| Mother smoked in pregnancy | No | 90.3 | 90.3 | 91.1 | 90.2 | 91.1 | 90.4 | 90.3 | 90.5 | 90.6 |
|  | Yes | 9.7 | 9.7 | 8.8 | 9.8 | 8.9 | 9.6 | 9.7 | 9.5 | 9.4 |
| Mother's depressive/anxiety symptoms in pregnancy | No | 90.1 | 90.5 | 89.2 | 90.3 | 90.7 | 90.6 | 90.5 | 90.7 | 90.8 |
|  | Yes | 9.9 | 9.5 | 9.1 | 9.7 | 9.3 | 9.4 | 9.5 | 9.3 | 9.2 |
|  | Missing | 0.0 | 0.0 | 1.7 | 0.0 | 0.0 | 0.0 | 0.0 | 0.0 | 0.0 |
| Mother has experienced physical/sexual violence | No | 73.8 | 74.1 | 72.2 | 73.9 | 74.9 | 74.4 | 74.1 | 74.6 | 75.1 |
|  | Yes | 26.2 | 25.9 | 24.2 | 26.1 | 25.1 | 25.6 | 25.9 | 25.4 | 24.9 |
|  | Missing | 0.0 | 0.0 | 3.6 | 0.0 | 0.0 | 0.0 | 0.0 | 0.0 | 0.0 |
| Unplanned pregnancy | No | 82.5 | 82.8 | 83.4 | 82.2 | 83.8 | 82.6 | 82.8 | 83.2 | 82.8 |
|  | Yes | 17.5 | 17.2 | 16.0 | 17.8 | 16.2 | 17.4 | 17.2 | 16.8 | 17.2 |
|  | Missing | 0.0 | 0.0 | 0.7 | 0.0 | 0.0 | 0.0 | 0.0 | 0.0 | 0.0 |
| Child's delayed language skills, 18 months | No | 71.6 | 72.4 | 71.7 | 71.4 | 71.5 | 72.7 | 72.4 | 73.7 | 73.1 |
|  | Yes | 3.2 | 3.2 | 3.2 | 3.2 | 3.2 | 3.0 | 3.2 | 3.0 | 3.1 |
|  | Missing | 25.3 | 24.4 | 25.0 | 25.4 | 25.3 | 24.3 | 24.4 | 23.3 | 23.8 |
| Child's delayed motor skills, 18 months | No | 70.8 | 71.6 | 70.9 | 70.6 | 70.7 | 71.9 | 71.6 | 72.9 | 72.3 |
|  | Yes | 3.3 | 3.4 | 3.3 | 3.4 | 3.3 | 3.1 | 3.4 | 3.1 | 3.2 |
|  | Missing | 25.9 | 25.0 | 25.7 | 26.0 | 26.0 | 25.0 | 25.0 | 24.0 | 24.5 |
| Child's delayed language skills, 36 months | No | 57.9 | 58.6 | 59.1 | 57.2 | 58.4 | 58.2 | 58.6 | 59.6 | 58.8 |
|  | Yes | 1.4 | 1.3 | 1.4 | 1.4 | 1.4 | 1.3 | 1.3 | 1.3 | 1.4 |
|  | Missing | 40.7 | 40.0 | 39.5 | 41.4 | 40.2 | 40.5 | 40.0 | 39.1 | 39.8 |
| Child's delayed motor skills, 36 months | No | 54.4 | 55.0 | 55.4 | 53.7 | 54.8 | 54.6 | 55.0 | 55.9 | 55.1 |
|  | Yes | 4.2 | 4.3 | 4.3 | 4.1 | 4.3 | 4.1 | 4.3 | 4.3 | 4.3 |
|  | Missing | 41.4 | 40.7 | 40.3 | 42.2 | 41.0 | 41.2 | 40.7 | 39.9 | 40.6 |

**Supplementary Table 3:** Details for multiple imputation, main analytic sample.

| **Supplementary Table 3: Details for multiple imputation, main analytic sample (N=68325):** |  |  |  |  |
| --- | --- | --- | --- | --- |
| **Imputed variable** | **n(missing)** | **n(cases)** | **Imputation model** | **Independent variables** |
| PAE week 1-15 (reported at Q1) | 4732 | 10,791 | logit | Child's sex and year of birth. Mother's and father's: age, age squared, educational qualifications, income. Mother's: number of previous pregnancies, smoking, depressive/anxiety symptoms, experience of physical/sexual violence, whether pregnancy was planned. PAE at week 13-30.<br>Auxiliary variables predicting PAE: weekly occasions, units per occasion, and frequency of binge drinking in 3 months prior to pregnancy. Other reports of PAE in any trimester. BMI, units of alcohol needed to feel an effect, social connectedness, life satisfaction, weekly exercise, as reported at Q1.<br>Child's ASQ outcomes and age when child first walked.<br>Child's English, reading and numeracy scores from grades 5, 8, and 9. |
| PAE week 13-30 (reported at Q3) | 8871 | 7,385 | logit | Child's sex and year of birth. Mother's and father's: age, age squared, educational qualifications, income. Mother's: number of previous pregnancies, smoking, depressive/anxiety symptoms, experience of physical/sexual violence, whether pregnancy was planned. PAE at week 1-15.<br>Auxiliary variables predicting PAE: weekly occasions, units per occasion, and frequency of binge drinking in 3 months prior to pregnancy. Other reports of PAE in any trimester. BMI, units of alcohol needed to feel an effect, social connectedness, life satisfaction, weekly exercise, as reported at Q1.<br>Child's ASQ outcomes and age when child first walked.<br>Child's English, reading and numeracy scores from grades 5, 8, and 9. |
| PAE week 1-13 (reported at Q3) | 10611 | 18,431 | logit | Child's sex and year of birth. Mother's and father's: age, age squared, educational qualifications, income. Mother's: number of previous pregnancies, smoking, depressive/anxiety symptoms, experience of physical/sexual violence, whether pregnancy was planned. PAE at week 1-15, PAE at week 13-30.<br>Auxiliary variables predicting PAE: weekly occasions, units per occasion, and frequency of binge drinking in 3 months prior to pregnancy. Other reports of PAE in any trimester. BMI, units of alcohol needed to feel an effect, social connectedness, life satisfaction, weekly exercise, as reported at Q1.<br>Child's ASQ outcomes and age when child first walked.<br>Child's English, reading and numeracy scores from grades 5, 8, and 9. |
| PAE reported at Q2 | 7725 | 6,725 | logit | Child's sex and year of birth. Mother's and father's: age, age squared, educational qualifications, income. Mother's: number of previous pregnancies, smoking, depressive/anxiety symptoms, experience of physical/sexual violence, whether pregnancy was planned. PAE at week 1-15, PAE at week 13-30.<br>Auxiliary variables predicting PAE: weekly occasions, units per occasion, and frequency of binge drinking in 3 months prior to pregnancy. Other reports of PAE<br>in any trimester. BMI, units of alcohol needed to feel an effect, social connectedness, life satisfaction, weekly exercise, as reported at Q1. Child's ASQ outcomes and age when child first walked. Child's English, reading and numeracy scores from grades 5, 8, and 9. |
| Child's delayed language skills, 18 months | 17277 | 2,161 | logit | Child's sex and year of birth. Mother's and father's: age, age squared, educational qualifications, income. Mother's: number of previous pregnancies, smoking, depressive/anxiety symptoms, experience of physical/sexual violence, whether pregnancy was planned. All PAE variables. Child's other ASQ outcomes and age when child first walked. Child's English, reading and numeracy scores from grades 5, 8, and 9. Child's exact age in months at completion of the questionnaire. |
| Child's delayed motor skills, 18 months | 17701 | 2,267 | logit | Child's sex and year of birth. Mother's and father's: age, age squared, educational qualifications, income. Mother's: number of previous pregnancies, smoking, depressive/anxiety symptoms, experience of physical/sexual violence, whether pregnancy was planned. All PAE variables. Child's other ASQ outcomes and age when child first walked. Child's English, reading and numeracy scores from grades 5, 8, and 9. Child's exact age in months at completion of the questionnaire. |
| Child's delayed language skills, 36 months | 27810 | 941 | logit | Child's sex and year of birth. Mother's and father's: age, age squared, educational qualifications, income. Mother's: number of previous pregnancies, smoking and depressive/anxiety symptoms, experience of physical/sexual violence. All PAE variables. Child's other ASQ outcomes and age when child first walked. Child's English, reading and numeracy scores from grades 5, 8, and 9. Child's exact age in months at completion of the questionnaire. |
| Child's delayed motor skills, 36 months | 28292 | 2,855 | logit | Child's sex and year of birth. Mother's and father's: age, age squared, educational qualifications, income. Mother's: number of previous pregnancies, smoking, depressive/anxiety symptoms, experience of physical/sexual violence, whether pregnancy was planned. All PAE variables. Child's other ASQ outcomes and age when child first walked. Child's English, reading and numeracy scores from grades 5, 8, and 9. Child's exact age in months at completion of the questionnaire. |
| Child's age at walking (months) | 13524 | N/A | predictive mean matching, knn(10) | Child's sex and year of birth. Mother's and father's: age, age squared, educational qualifications, income. Mother's: number of previous pregnancies, smoking, depressive/anxiety symptoms, experience of physical/sexual violence, whether pregnancy was planned. All PAE variables. Child's ASQ outcomes. |
|  |  |  |  | Child's English, reading and numeracy scores from grades 5, 8, and 9. |
| Notes: for stratified and additional analyses, separate imputation models were run as above but with the following variations: for genotype-stratified effects, imputation models were run separately for each stratum of maternal and child genotype. For sensitivity analyses examining effects in first-born children only and within specific time periods, duos were dropped as required before imputation. For sensitivity analyses examining categorical PAE in week 1-13 (none/some/binge), a three-group categorical was imputed in place of the binary variable. |  |  |  |  |

**Supplementary Table 4:**
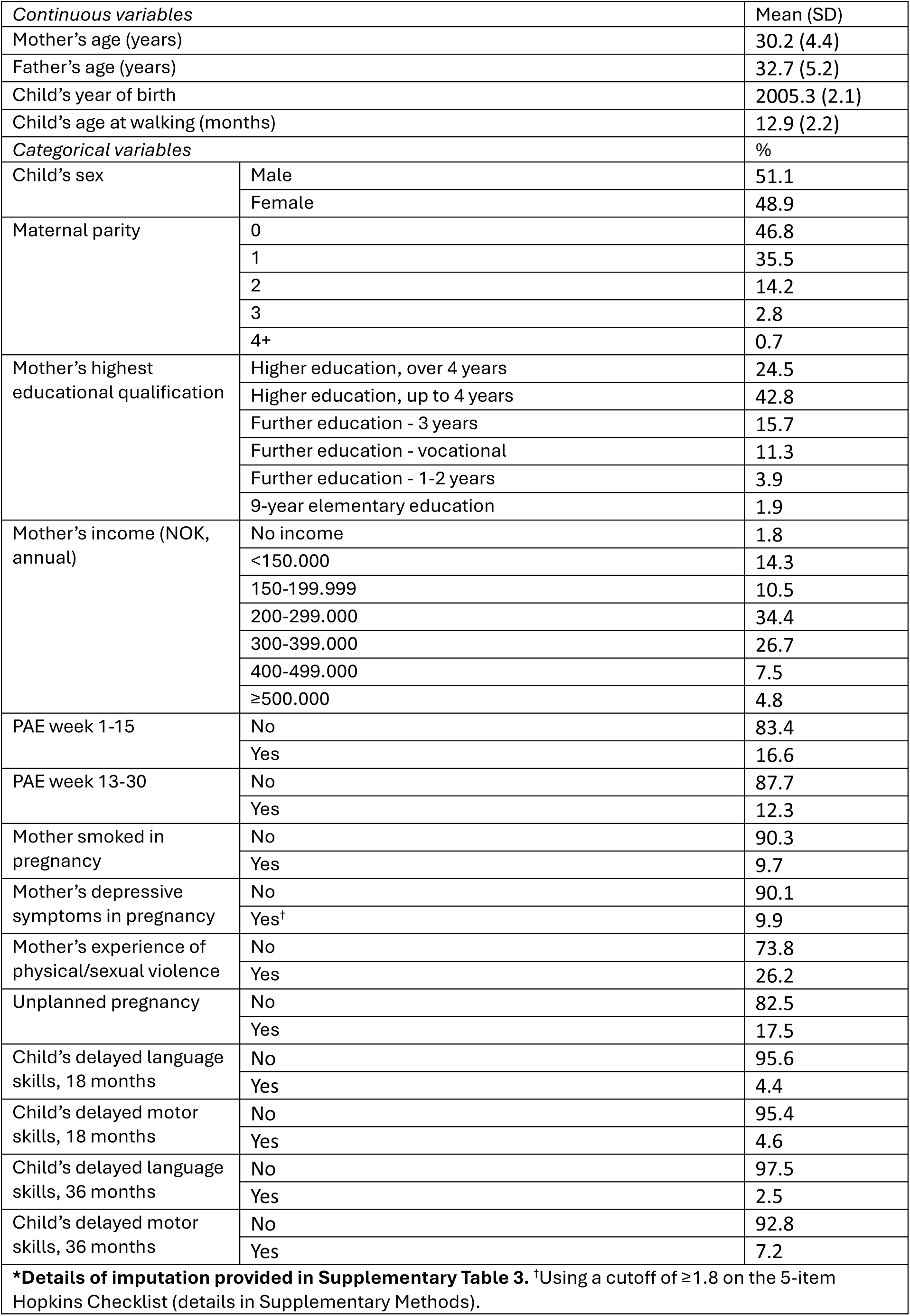
Descriptive characteristics of analytic sample for main analysis (post-imputation)

**Supplementary Table 5:** Secular and socioeconomic patterning of PAE.

| Supplementary Table 5: Secular and socioeconomic patterning of PAE* |  |  |  |  |  |
| --- | --- | --- | --- | --- | --- |
|  |  | PAE week 1-15 | PAE week 13-30 | PAE week 1-15:<br>'some'<sup>†</sup> | PAE week 1-15:<br>'binge'<sup>†</sup> |
|  |  | OR (95%CI) | OR (95%CI) | OR (95%CI) | OR (95%CI) |
| Child's year of birth | Change per year | 0.83 (0.82,0.84) | 0.72 (0.71,0.73) | 0.76 (0.75,0.77) | 0.96 (0.94,0.98) |
| Mother's highest educational qualification<br>(Reference: higher education, over 4 years) | Higher education, up to 4 years | 0.85 (0.80,0.90) | 0.83 (0.77,0.88) | 0.78 (0.72,0.83) | 0.99 (0.91,1.08) |
|  | Further education - 3 years | 0.89 (0.82,0.96) | 0.87 (0.80,0.96) | 0.82 (0.74,0.90) | 1.06 (0.94,1.19) |
|  | Further education - vocational | 0.74 (0.68,0.81) | 0.70 (0.63,0.79) | 0.70 (0.63,0.79) | 0.85 (0.74,0.98) |
|  | Further education - 1-2 years | 0.70 (0.61,0.80) | 0.72 (0.61,0.85) | 0.62 (0.52,0.74) | 0.89 (0.73,1.10) |
|  | 9-year elementary education | 0.85 (0.71,1.02) | 0.8 (0.63,1.01) | 0.83 (0.66,1.05) | 0.97 (0.73,1.28) |
| Father's highest educational qualification<br>(Reference: higher education, over 4 years) | Higher education, up to 4 years | 0.97 (0.92,1.03) | 0.89 (0.83,0.96) | 0.93 (0.87,1.01) | 1.07 (0.97,1.17) |
|  | Further education - 3 years | 0.97 (0.90,1.05) | 0.85 (0.78,0.93) | 0.90 (0.82,0.99) | 1.12 (1.00,1.26) |
|  | Further education - vocational | 0.86 (0.80,0.93) | 0.73 (0.67,0.79) | 0.75 (0.69,0.82) | 1.07 (0.96,1.19) |
|  | Further education - 1-2 years | 0.91 (0.81,1.02) | 0.74 (0.64,0.85) | 0.78 (0.68,0.90) | 1.18 (1.00,1.40) |
|  | 9-year elementary education | 0.87 (0.77,0.98) | 0.75 (0.64,0.87) | 0.75 (0.64,0.88) | 1.10 (0.91,1.32) |
| Mother's income (NOK, annual)<br>(Reference: ≥500.000) | 400-499.000 | 0.87 (0.78,0.97) | 0.83 (0.73,0.94) | 0.81 (0.71,0.92) | 0.98 (0.82,1.17) |
|  | 300-399.000 | 0.78 (0.71,0.85) | 0.68 (0.61,0.76) | 0.7 (0.63,0.78) | 0.91 (0.78,1.06) |
|  | 200-299.000 | 0.65 (0.59,0.71) | 0.54 (0.48,0.60) | 0.58 (0.52,0.65) | 0.76 (0.65,0.89) |
|  | 150-199.999 | 0.66 (0.59,0.75) | 0.58 (0.50,0.66) | 0.59 (0.51,0.68) | 0.8 (0.66,0.96) |
|  | <150.000 | 0.73 (0.65,0.82) | 0.56 (0.49,0.64) | 0.65 (0.57,0.75) | 0.88 (0.73,1.05) |
|  | No income | 0.71 (0.59,0.86) | 0.67 (0.54,0.84) | 0.67 (0.53,0.84) | 0.76 (0.56,1.03) |
| Father's income (NOK, annual)<br>(Reference: ≥500.000) | 400-499.000 | 0.88 (0.82,0.94) | 0.87 (0.80,0.94) | 0.90 (0.82,0.98) | 0.87 (0.78,0.97) |
|  | 300-399.000 | 0.92 (0.86,0.98) | 0.85 (0.79,0.91) | 0.92 (0.85,0.99) | 0.93 (0.84,1.03) |
|  | 200-299.000 | 0.90 (0.83,0.97) | 0.84 (0.77,0.92) | 0.89 (0.81,0.98) | 0.91 (0.81,1.02) |
|  | 150-199.999 | 1.00 (0.88,1.13) | 0.96 (0.82,1.11) | 1.03 (0.88,1.21) | 0.97 (0.80,1.16) |
|  | <150.000 | 1.14 (1.02,1.28) | 1.06 (0.92,1.22) | 1.22 (1.06,1.41) | 1.05 (0.89,1.23) |
|  | No income | 1.18 (0.93,1.50) | 1.12 (0.84,1.51) | 1.36 (1.01,1.84) | 1.01 (0.70,1.45) |
| *All associations mutually adjusted. Also adjusted for: mother's and father's age and age squared, and mother's number of previous pregnancies. <sup>†</sup>From a multinomial logistic model with reference outcome of no drinking at week 1-15. 'Some' defined as >0 but <5 units on any given occasion, 'binge' as 5+ units on at least one occasion. |  |  |  |  |  |

**Supplementary Table 6:** Fully-adjusted associations of prenatal alcohol exposure (PAE) and all developmental outcomes, main and alternative specifications.

| Supplementary Table 6: Fully adjusted associations of prenatal alcohol exposure (PAE) and all developmental outcomes in the whole sample, main and alternative specifications |  |  |  |  |  |  |  |
| --- | --- | --- | --- | --- | --- | --- | --- |
|  |  | Main specification | First-born children only | Birth year: 2000-2005 | Birth year: 2006-2009 | Categorical PAE |  |
|  | Exposure period | OR (95%CI) | OR (95%CI) | OR (95%CI) | OR (95%CI) | Heaviness | OR (95%CI) |
| Delayed language skills at 18 months* | Week 0-15 | 1.14 (0.99,1.31) | 1.21 (0.99,1.47) | 1.09 (0.91,1.31) | 1.22 (1.00,1.49) | Some<sup>‡</sup> | 1.22 (1.03,1.43) |
|  | Week 13-30 | 1.06 (0.92,1.23) | 1.18 (0.92,1.50) | 0.98 (0.79,1.21) | 1.24 (0.97,1.58) | Binge<sup>‡</sup> | 1.06 (0.88,1.27) |
| Delayed motor skills at 18 months* | Week 0-15 | 0.95 (0.84,1.07) | 0.89 (0.71,1.10) | 1.00 (0.84,1.19) | 0.89 (0.73,1.10) | Some<sup>‡</sup> | 0.98 (0.83,1.15) |
|  | Week 13-30 | 0.99 (0.85,1.15) | 0.93 (0.71,1.22) | 0.95 (0.79,1.15) | 1.05 (0.80,1.37) | Binge<sup>‡</sup> | 0.93 (0.75,1.14) |
| Delayed language skills at 36 months* | Week 0-15 | 1.09 (0.89,1.34) | 0.96 (0.69,1.32) | 1.07 (0.81,1.40) | 1.13 (0.82,1.56) | Some<sup>‡</sup> | 1.05 (0.82,1.35) |
|  | Week 13-30 | 0.96 (0.78,1.18) | 1.25 (0.84,1.87) | 0.92 (0.70,1.22) | 1.06 (0.70,1.62) | Binge<sup>‡</sup> | 1.12 (0.85,1.48) |
| Delayed motor skill at 36 months* | Week 0-15 | 1.06 (0.93,1.21) | 1.14 (0.95,1.36) | 1.06 (0.89,1.26) | 1.06 (0.89,1.26) | Some<sup>‡</sup> | 1.00 (0.86,1.16) |
|  | Week 13-30 | 1.00 (0.86,1.17) | 0.98 (0.80,1.21) | 1.00 (0.82,1.21) | 0.98 (0.78,1.24) | Binge<sup>‡</sup> | 1.13 (0.96,1.33) |
|  |  | Mean difference (95%CI) | Mean difference (95%CI) | Mean difference (95%CI) | Mean difference (95%CI) | Heaviness | Mean difference (95%CI) |
| Age at walking (months)<sup>†</sup> | Week 0-15 | -0.04 (-0.10,0.02) | -0.03 (-0.12,0.05) | -0.05 (-0.13,0.03) | -0.02 (-0.11,0.07) | Some<sup>‡</sup> | -0.02 (-0.10,0.06) |
|  | Week 13-30 | 0.01 (-0.07,0.08) | 0.00 (-0.11,0.12) | 0.02 (-0.07,0.11) | 0.00 (-0.12,0.12) | Binge<sup>‡</sup> | -0.06 (-0.14,0.01) |
| *Adjusted for child's: sex, year of birth, and exact age in months at questionnaire completion, mother's and father's: age, age squared, number of previous pregnancies, highest educational qualification, and income in pregnancy, mother's: number of previous pregnancies, smoking and depressive/anxiety symptoms in pregnancy, experience of physical/sexual violence, whether the pregnancy was planned. <sup>†</sup>Adjusted for child's: sex, year of birth, mother's and father's: age, age squared, number of previous pregnancies, highest educational qualification, and income in pregnancy, mother's: number of previous pregnancies, smoking and depressive/anxiety symptoms in pregnancy, experience of physical/sexual violence, whether the pregnancy was planned. <sup>‡</sup>At week 1-15 (lower drinking prevalence at week 13-30 precluded subgroup analysis for this period). |  |  |  |  |  |  |  |

**Supplementary Table 7:**
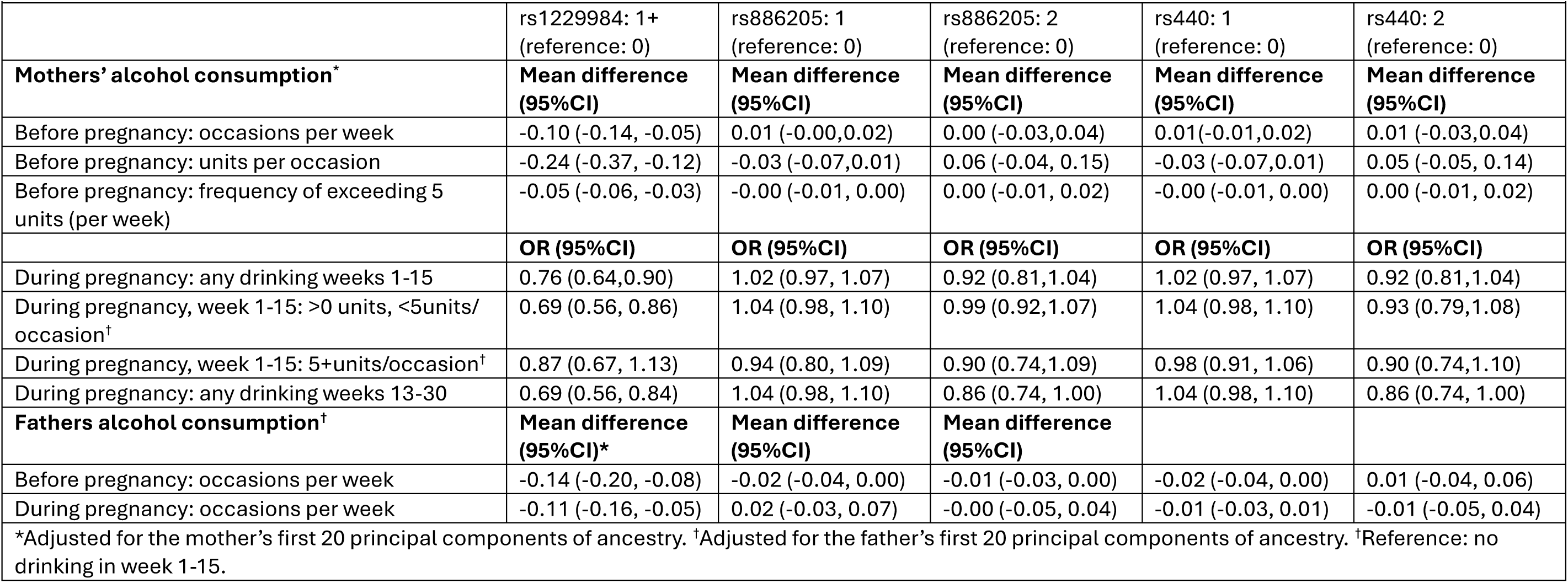
Associations of mother’s and father’s ADH1B and ALDH2 genotypes with alcohol consumption.

|  |  |  |  |  |  |
| --- | --- | --- | --- | --- | --- |
|  | rs1229984: 1+<br>(reference: 0) | rs886205: 1<br>(reference: 0) | rs886205: 2<br>(reference: 0) | rs440: 1<br>(reference: 0) | rs440: 2<br>(reference: 0) |
| **Mothers' alcohol consumption\*** | **Mean difference**<br>**(95%CI)** | **Mean difference**<br>**(95%CI)** | **Mean difference**<br>**(95%CI)** | **Mean difference**<br>**(95%CI)** | **Mean difference**<br>**(95%CI)** |
| Before pregnancy: occasions per week | -0.10 (-0.14, -0.05) | 0.01 (-0.00, 0.02) | 0.00 (-0.03, 0.04) | 0.01 (-0.01, 0.02) | 0.01 (-0.03, 0.04) |
| Before pregnancy: units per occasion | -0.24 (-0.37, -0.12) | -0.03 (-0.07, 0.01) | 0.06 (-0.04, 0.15) | -0.03 (-0.07, 0.01) | 0.05 (-0.05, 0.14) |
| Before pregnancy: frequency of exceeding 5<br>units (per week) | -0.05 (-0.06, -0.03) | -0.00 (-0.01, 0.00) | 0.00 (-0.01, 0.02) | -0.00 (-0.01, 0.00) | 0.00 (-0.01, 0.02) |
|  | **OR (95%CI)** | **OR (95%CI)** | **OR (95%CI)** | **OR (95%CI)** | **OR (95%CI)** |
| During pregnancy: any drinking weeks 1-15 | 0.76 (0.64, 0.90) | 1.02 (0.97, 1.07) | 0.92 (0.81, 1.04) | 1.02 (0.97, 1.07) | 0.92 (0.81, 1.04) |
| During pregnancy, week 1-15: >0 units, <5units/<br>occasion<sup>†</sup> | 0.69 (0.56, 0.86) | 1.04 (0.98, 1.10) | 0.99 (0.92, 1.07) | 1.04 (0.98, 1.10) | 0.93 (0.79, 1.08) |
| During pregnancy, week 1-15: 5+units/occasion<sup>†</sup> | 0.87 (0.67, 1.13) | 0.94 (0.80, 1.09) | 0.90 (0.74, 1.09) | 0.98 (0.91, 1.06) | 0.90 (0.74, 1.10) |
| During pregnancy: any drinking weeks 13-30 | 0.69 (0.56, 0.84) | 1.04 (0.98, 1.10) | 0.86 (0.74, 1.00) | 1.04 (0.98, 1.10) | 0.86 (0.74, 1.00) |
| **Fathers alcohol consumption<sup>†</sup>** | **Mean difference**<br>**(95%CI)\*** | **Mean difference**<br>**(95%CI)** | **Mean difference**<br>**(95%CI)** |  |  |
| Before pregnancy: occasions per week | -0.14 (-0.20, -0.08) | -0.02 (-0.04, 0.00) | -0.01 (-0.03, 0.00) | -0.02 (-0.04, 0.00) | 0.01 (-0.04, 0.06) |
| During pregnancy: occasions per week | -0.11 (-0.16, -0.05) | 0.02 (-0.03, 0.07) | -0.00 (-0.05, 0.04) | -0.01 (-0.03, 0.01) | -0.01 (-0.05, 0.04) |
| *Adjusted for the mother's first 20 principal components of ancestry. <sup>†</sup>Adjusted for the father's first 20 principal components of ancestry. <sup>†</sup>Reference: no drinking in week 1-15. |  |  |  |  |  |

**Supplementary Table 8:**
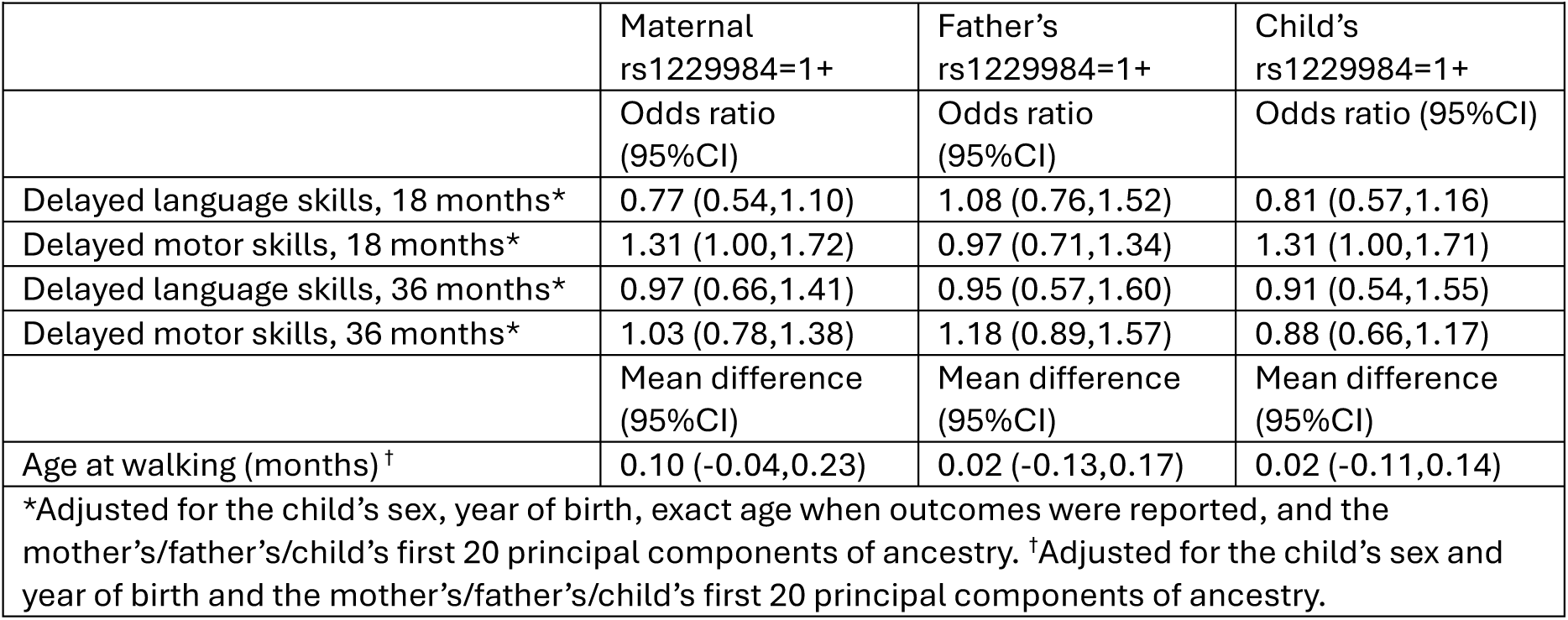
Association of maternal, paternal, and child’s own rs1229984 genotype with all children’s outcomes.

**Supplementary Table 9:** N cases for imputed outcomes in genotype-stratified groups.

|  |  |  |  |  |  |
| --- | --- | --- | --- | --- | --- |
| **Imputed variable** | **N cases:**<br>**maternal**<br>**rs1229984=0** | **N cases:**<br>**maternal**<br>**rs1229984=1+** | **N cases:**<br>**child**<br>**rs1229984=0** | **N cases:**<br>**child**<br>**rs1229984=1+** | **N predictor**<br>**variables in**<br>**imputation**<br>**model\*** |
| Child's delayed language skills, 18 months | 1,805 | 34 | 1,465 | 31 | 31 |
| Child's delayed motor skills, 18 months | 1,903 | 53 | 1,513 | 45 | 31 |
| Child's delayed language skills, 36 months | 755 | 17 | 627 | 14 | 31 |
| Child's delayed motor skills, 36 months | 2,416 | 57 | 2,071 | 47 | 31 |
| *Imputation models for genotype-stratified analyses were specified as in Supplementary Table 2 but run separately for each stratum of maternal and child rs1229984. |  |  |  |  |  |

**Supplementary Table 10:** Mutually-adjusted associations of mother’s and child’s own rs1229984 genotypes with all outcomes, stratified by PAE at week 1-15.

| **Supplementary Table 10: Mutually-adjusted associations of mother's and children's rs1229984 genotypes with all outcomes, stratified by PAE at week 1-15** |  |  |  |  |
| --- | --- | --- | --- | --- |
|  | **PAE at week-15: no** |  | **PAE at week-15: yes** |  |
|  | **Maternal rs1229984\*** | **Child's rs1229984\*** | **Maternal rs1229984\*** | **Child's rs1229984\*** |
|  | Odds ratio (95%CI) | Odds ratio (95%CI) | Odds ratio (95%CI) | Odds ratio (95%CI) |
| Delayed language skills, 18 months* | 0.74 (0.44,1.26) | 0.94 (0.58,1.53) | 0.64 (0.17,2.49) | 0.76 (0.24,2.41) |
| Delayed motor skills, 18 months* | 1.14 (0.75,1.72) | 1.22 (0.83,1.80) | 1.09 (0.33,3.64) | 1.61 (0.63,4.14) |
| Delayed language skills, 36 months* | 0.83 (0.43,1.61) | 0.99 (0.55,1.81) | 3.34 (1.09,10.18) | 0.18 (0.02,1.41) |
| Delayed motor skills, 36 months* | 1.02 (0.70,1.49) | 0.97 (0.67,1.41) | 1.25 (0.46,3.44) | 0.34 (0.09,1.29) |
|  | Mean difference (95%CI) | Mean difference (95%CI) | Mean difference (95%CI) | Mean difference (95%CI) |
| Age at walking (months)<sup>†</sup> | 0.06 (-0.12,0.24) | -0.07 (-0.24,0.10) | 0.16 (-0.31,0.63) | 0.30 (-0.16,0.76) |
| *Reference: 0 effect alleles (acetaldehyde-increasing alleles). Effects of maternal and child genotype are adjusted for each other. Also adjusted for: the child's sex, year of birth, exact age when outcomes were reported, and the mother's and child's first 20 principal components of ancestry. <sup>†</sup>Effects of maternal and child genotype are adjusted for each other. Also adjusted for: the child's sex and year of birth and the mother's and child's first 20 principal components of ancestry. |  |  |  |  |

**Supplementary Table 11:** Adjusted associations of prenatal alcohol exposure (PAE) and all developmental outcomes, stratified by maternal and child rs1229984 genotype.

| **Supplementary Table 11: Fully adjusted associations of prenatal alcohol exposure (PAE) and all developmental outcomes, stratified by maternal and child rs1229984 genotype** |  |  |  |  |  |  |
| --- | --- | --- | --- | --- | --- | --- |
|  |  | All genotype groups | Maternal rs1229984=0 | Maternal rs1229984=1+ | Child's rs1229984=0 | Child's rs1229984=1+ |
|  | Exposure period | OR (95%CI) | Odds ratio (95%CI) | Odds ratio (95%CI) | Odds ratio (95%CI) | Odds ratio (95%CI) |
| Delayed language skills, 18 months* | Week 0-15 | 1.14 (0.99,1.31) | 1.16 (1.01,1.35) | 1.90 (0.64,5.62) | 1.15 (0.98,1.34) | 1.75 (0.57,5.39) |
|  | Week 13-30 | 1.06 (0.92,1.23) | 1.09 (0.91,1.29) | 0.63 (0.12,3.41) | 1.13 (0.93,1.36) | 0.29 (0.05,1.75) |
| Delayed motor skills, 18 months* | Week 0-15 | 0.95 (0.84,1.07) | 0.90 (0.77,1.05) | 1.04 (0.36,3.00) | 0.97 (0.83,1.13) | 1.17 (0.44,3.09) |
|  | Week 13-30 | 0.99 (0.85,1.15) | 1.03 (0.87,1.23) | 0.64 (0.13,3.17) | 0.95 (0.77,1.16) | 0.97 (0.30,3.13) |
| Delayed language skills, 36 months* | Week 0-15 | 1.09 (0.89,1.34) | 1.08 (0.85,1.38) | 2.80 (0.55,14.24) | 1.08 (0.85,1.37) | 0.62 (0.07,5.42) |
|  | Week 13-30 | 0.96 (0.78,1.18) | 0.99 (0.76,1.30) | 0.46 (0.05,4.06) | 1.01 (0.78,1.32) | 1.02 (0.10,10.26) |
| Delayed motor skills, 36 months* | Week 0-15 | 1.06 (0.93, 1.21) | 1.05 (0.92,1.21) | 1.25 (0.54,2.90) | 1.14 (0.98,1.33) | 0.42 (0.11,1.55) |
|  | Week 13-30 | 1.00 (0.86,1.17) | 1.03 (0.86,1.23) | 0.44 (0.12,1.59) | 1.03 (0.88,1.22) | 1.44 (0.45,4.64) |
|  |  | Mean difference (95%CI) | Mean difference (95%CI) | Mean difference (95%CI) | Mean difference (95%CI) | Mean difference (95%CI) |
| Age at walking (months)<sup>†</sup> | Week 0-15 | -0.04 (-0.10,0.02) | -0.05 (-0.12,0.02) | 0.20 (-0.24,0.65) | -0.02 (-0.09,0.05) | 0.21 (-0.21,0.63) |
|  | Week 13-30 | -0.01 (-0.07,0.08) | 0.03 (-0.05,0.11) | -0.14 (-0.63,0.34) | 0.01 (-0.07,0.09) | 0.10 (-0.39,0.58) |
| *Adjusted for child's: sex, year of birth, and exact age in months at questionnaire completion, mother's and father's: age, age squared, number of previous pregnancies, highest educational qualification, and income in pregnancy, mother's: number of previous pregnancies, smoking and depressive/anxiety symptoms in pregnancy, experience of physical/sexual violence, whether the pregnancy was planned. <sup>†</sup>Adjusted for child's: sex, year of birth, mother's and father's: age, age squared, number of previous pregnancies, highest educational qualification, and income in pregnancy, mother's: number of previous pregnancies, smoking and depressive/anxiety symptoms in pregnancy, experience of physical/sexual violence, whether the pregnancy was planned. |  |  |  |  |  |  |

**Supplementary Table 12:**
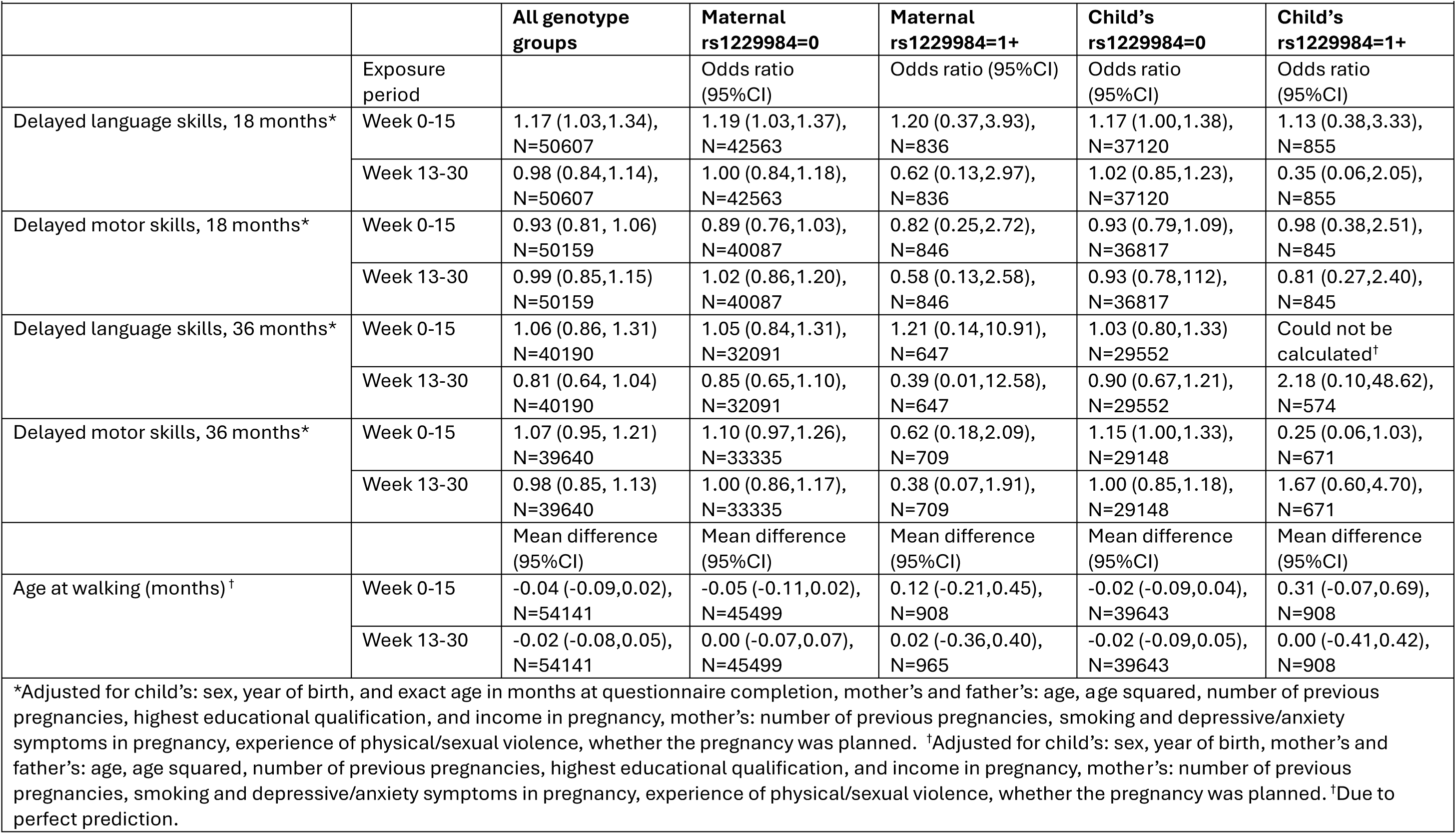
Complete-case analysis: adjusted associations of prenatal alcohol exposure (PAE) and all developmental outcomes in the whole sample and by genotype.

|  |  |  |  |  |  |  |
| --- | --- | --- | --- | --- | --- | --- |
|  |  | **All genotype groups** | **Maternal rs1229984=0** | **Maternal rs1229984=1+** | **Child's rs1229984=0** | **Child's rs1229984=1+** |
|  | Exposure period |  | Odds ratio (95%CI) | Odds ratio (95%CI) | Odds ratio (95%CI) | Odds ratio (95%CI) |
| Delayed language skills, 18 months* | Week 0-15 | 1.17 (1.03,1.34), N=50607 | 1.19 (1.03,1.37), N=42563 | 1.20 (0.37,3.93), N=836 | 1.17 (1.00,1.38), N=37120 | 1.13 (0.38,3.33), N=855 |
|  | Week 13-30 | 0.98 (0.84,1.14), N=50607 | 1.00 (0.84,1.18), N=42563 | 0.62 (0.13,2.97), N=836 | 1.02 (0.85,1.23), N=37120 | 0.35 (0.06,2.05), N=855 |
| Delayed motor skills, 18 months* | Week 0-15 | 0.93 (0.81, 1.06) N=50159 | 0.89 (0.76,1.03), N=40087 | 0.82 (0.25,2.72), N=846 | 0.93 (0.79,1.09), N=36817 | 0.98 (0.38,2.51), N=845 |
|  | Week 13-30 | 0.99 (0.85,1.15) N=50159 | 1.02 (0.86,1.20), N=40087 | 0.58 (0.13,2.58), N=846 | 0.93 (0.78,1.12), N=36817 | 0.81 (0.27,2.40), N=845 |
| Delayed language skills, 36 months* | Week 0-15 | 1.06 (0.86, 1.31) N=40190 | 1.05 (0.84,1.31), N=32091 | 1.21 (0.14,10.91), N=647 | 1.03 (0.80,1.33) N=29552 | Could not be calculated<sup>†</sup> |
|  | Week 13-30 | 0.81 (0.64, 1.04) N=40190 | 0.85 (0.65,1.10), N=32091 | 0.39 (0.01,12.58), N=647 | 0.90 (0.67,1.21), N=29552 | 2.18 (0.10,48.62), N=574 |
| Delayed motor skills, 36 months* | Week 0-15 | 1.07 (0.95, 1.21) N=39640 | 1.10 (0.97,1.26), N=33335 | 0.62 (0.18,2.09), N=709 | 1.15 (1.00,1.33), N=29148 | 0.25 (0.06,1.03), N=671 |
|  | Week 13-30 | 0.98 (0.85, 1.13) N=39640 | 1.00 (0.86,1.17), N=33335 | 0.38 (0.07,1.91), N=709 | 1.00 (0.85,1.18), N=29148 | 1.67 (0.60,4.70), N=671 |
|  |  | Mean difference (95%CI) | Mean difference (95%CI) | Mean difference (95%CI) | Mean difference (95%CI) | Mean difference (95%CI) |
| Age at walking (months)<sup>†</sup> | Week 0-15 | -0.04 (-0.09,0.02), N=54141 | -0.05 (-0.11,0.02), N=45499 | 0.12 (-0.21,0.45), N=908 | -0.02 (-0.09,0.04), N=39643 | 0.31 (-0.07,0.69), N=908 |
|  | Week 13-30 | -0.02 (-0.08,0.05), N=54141 | 0.00 (-0.07,0.07), N=45499 | 0.02 (-0.36,0.40), N=965 | -0.02 (-0.09,0.05), N=39643 | 0.00 (-0.41,0.42), N=908 |
| *Adjusted for child's: sex, year of birth, and exact age in months at questionnaire completion, mother's and father's: age, age squared, number of previous pregnancies, highest educational qualification, and income in pregnancy, mother's: number of previous pregnancies, smoking and depressive/anxiety symptoms in pregnancy, experience of physical/sexual violence, whether the pregnancy was planned. <sup>†</sup>Adjusted for child's: sex, year of birth, mother's and father's: age, age squared, number of previous pregnancies, highest educational qualification, and income in pregnancy, mother's: number of previous pregnancies, smoking and depressive/anxiety symptoms in pregnancy, experience of physical/sexual violence, whether the pregnancy was planned. <sup>†</sup>Due to perfect prediction. |  |  |  |  |  |  |

